# Hepatic Fibro-Inflammation and Atrial Fibrillation: A Dual-Track Metabolic Axis Revealed by a Metabolomic Clock

**DOI:** 10.64898/2026.06.17.26354669

**Authors:** Guanyu Lin, Jinzhu Hu, Tongsheng Huang, Wenli Gu, Jingfeng Wang, Yiyang Cao, Linghua Fu, Zhaoyu Liu, Weiwen Lim, Ching Chi-Keong, Chrishan Ramachandra, Hualin Fan, Yinhua Zhang, Shuai Wei, Haifeng Zhang, Yuan Jiang, Yuling Zhang, Lin Zhang, Wengen Zhu, Peng Yu, Yangxin Chen, Xiao Liu, Derek John Hausenloy

## Abstract

**Aims:** Fatty liver disease has been associated with atrial fibrillation (AF), yet the liver-heart axis, the interplay between hepatic fibro-inflammation, systemic metabolism, and genetic susceptibility, remains poorly defined. We aimed to characterize this axis and its association with incident AF.

**Methods and Results:** In this prospective cohort study, liver fibrosis was assessed via four biochemical indices and magnetic resonance imaging (corrected T1 [cT1]). We integrated metabolome-wide causal mediation (249 nuclear magnetic resonance [NMR] features) with Elastic Net modelling, cardiac phenomapping (cardiac magnetic resonance and electrocardiogram), and unsupervised clustering. A metabolomic risk score (MRS) was derived and evaluated for gene-environment interactions with an AF polygenic risk score (PRS) and for incremental prediction beyond CHARGE-AF, ARIC, and C2HEST.

Among 403,974 UK Biobank participants (median follow-up 13.18 years), 26,677 developed incident AF. High-risk NAFLD fibrosis score (NFS; HR 1.54, 95% CI 1.43–1.66), Fibrosis-4 index (FIB-4; HR 1.53, 95% CI 1.44–1.62), and liver MRI cT1 (HR 1.41, 95% CI 1.11–1.79) were independently associated with AF. Phenomapping identified a dual-track axis: (1) systemic inflammation and lipotoxicity linked to electrophysiological alterations without chamber dilation, and (2) fatty-acid imbalance associated with structural enlargement. Three metabolomic clusters emerged; a “Fibro-Inflammatory” phenotype exhibited distinct metabolomic derangements, ketogenic stress, and a high residual AF risk independent of traditional comorbidities. The MRS compounded AF risk across all PRS strata and improved prediction beyond CHARGE-AF (ΔAUC +0.005; cNRI 10.2%), ARIC (ΔAUC +0.006; cNRI 11.7%), and C2HEST (ΔAUC +0.042; cNRI 32.2%).

**Conclusions:** Liver fibrosis is a robust predictor of AF. A fibro-inflammatory hepatic-metabolomic signature defines a modifiable axis that potentiates genetic susceptibility and enhances AF risk stratification. Targeting liver-derived metabolic dysfunction may offer a new therapeutic avenue for AF prevention.

**Translational Perspective:** Atrial fibrillation often arises outside conventional clinical and genetic risk tiers. Hepatic fibro-inflammation leaves a circulating metabolomic signature tracking distinct electrical and structural cardiac remodelling pathways, defining a lean, normolipidemic “Fibro-Inflammatory” phenotype carrying high residual AF risk that legacy scores systematically under-classify. A metabolomic risk score unmasked polygenic vulnerability and improved reclassification beyond CHARGE-AF, ARIC, and C2HEST. These findings position liver-derived metabolic dysregulation as a potentially modifiable axis for AF stratification—complementing clinical and genetic information and nominating candidate targets (atherogenic lipoproteins, systemic inflammation, ketogenic stress) for early prevention.

## 1. Introduction

Atrial fibrillation (AF) represents a growing global cause of cardiovascular morbidity and mortality, with rising incidence and clinical burden despite advances in prevention and management ^1^. Contemporary risk stratification relies primarily on clinical algorithms that aggregate historical end-organ damage and a polygenic risk score (PRS) that quantifies inherited susceptibility ^2^. Yet a considerable proportion of incident AF occurs in individuals without extreme genetic or clinical risk, underscoring the presence of dynamic, acquired “residual risk” that may shape subclinical arrhythmogenic substrates well before overt disease manifestation.

Metabolic dysfunction-associated steatotic liver disease (MASLD) and advanced hepatic fibro-inflammation have recently emerged as systemic conditions linked to heightened cardiovascular risk ^3–5^. Traditionally, the arrhythmogenic consequences of hepatic dysfunction have been attributed to haemodynamic perturbations and progressive left atrial enlargement. However, this paradigm incompletely captures the complex liver-heart biology. The specific subclinical pathways, circulating mediators, and metabolic signatures through which hepatic fibro-inflammation influences the heart remain incompletely understood ^6^. Accumulating evidence suggests that liver-derived metabolic and inflammatory stressors may promote arrhythmogenic remodelling beyond macroscopic chamber dilation, encompassing distinct electrophysiological and structural trajectories ^7^.

Genetic susceptibility and metabolic dysfunction each contribute to AF risk, yet their combined interplay is poorly defined. We hypothesize that a “double-hit” model, wherein acquired metabolic dysregulation compounds underlying polygenic vulnerability, may better explain interindividual variation in AF susceptibility. High-resolution metabolic profiling, including a metabolomic risk score (MRS) that functions as a virtual “metabolomic clock” of systemic health, offers a means to quantify this modifiable systemic state and refine early AF prevention ^8^.

Leveraging the depth of the UK Biobank cohort, we sought to delineate the trajectory from hepatic fibro-inflammation to incident AF. Specifically, we sought to: (1) characterize the independent association between hepatic fibro-inflammation and AF; (2) identify metabolic mediators and target-organ phenotypes underlying this liver-heart axis; and (3) derive an MRS to evaluate its interaction with polygenic susceptibility and its incremental predictive value beyond established clinical and genetic models.

## 2. Methods

### 2.1 Study Population and Design

The UK Biobank is a prospective cohort of approximately 500,000 participants aged 40-69 years recruited between 2006 and 2010 ^9^. All participants provided written informed consent, and the study received ethics approval from the North West Multicentre Research Ethics Committee (approval no. 11/NW/0382), and the investigation conforms to the principles outlined in the Declaration of Helsinki. We excluded participants with baseline AF or atrial flutter, missing data for AF PRS, genetic principal components (PCs) ^10^, or incomplete liver fibrosis indices, yielding a primary analytical cohort of 403,974 participants.

### 2.2 Ascertainment of Exposure and Outcome

For the primary analysis, liver fibrosis risk was assessed using four validated noninvasive tests: Fibrosis-4 (FIB-4) index, nonalcoholic fatty liver disease (NAFLD) fibrosis score (NFS), aspartate aminotransferase to platelet ratio index (APRI), and aspartate aminotransferase (AST) to alanine aminotransferase (ALT) ratio. Low-risk fibrosis was defined as a FIB-4 < 1.3 ^11^, NFS < −1.455 ^12^, APRI < 0.5 ^13^, or AST/ALT < 1 ^14^; high-risk fibrosis was defined as FIB-4 > 2.67, NFS > 0.675, APRI ≥ 0.5, or AST/ALT > 2. Participants who did not meet these criteria were classified into the ‘intermediate’ risk group.

In participants with valid corrected T1 (cT1) mapping and proton density fat fraction (PDFF) measurements, liver fibrosis was assessed using cT1, reflecting fibrosis and fluid retention ^15^. High-risk hepatic fibro-inflammation was defined as cT1 ≥ 750 ms, while low-risk was defined as cT1 < 750 ms.

The primary outcome of incident AF (ICD-10 code I48) was ascertained via hospital admissions and death registries. Follow-up extended from baseline to first AF diagnosis, death, loss to follow-up, or May 31, 2022.

### 2.3 Covariates, Multi-omics, and Phenotypic Data Acquisition

Baseline sociodemographic and lifestyle covariates included age, sex, ethnicity, Townsend Deprivation Index (TDI) ^16^, smoking status, alcohol intake, and body mass index (BMI). Baseline clinical comorbidities, including type 2 diabetes (T2DM), hypertension, dyslipidemia, chronic kidney disease (CKD), and cardiovascular disease (CVD), were defined based on ICD-9/10 codes, self-reported diagnoses, medication records, and laboratory biomarkers. Detailed laboratory measurement protocols, mathematical formulas for these indices, and definitions for covariates are provided in the **Supplementary Methods**. Missing data were handled using multiple imputation by chained equations (MICE) with predictive mean matching ^17^, with variable-specific missing rates detailed in **Supplementary Table S13**.

Genetic predisposition was assessed using a standardized AF PRS derived from a large-scale genome-wide association study (GWAS) meta-analysis ^18^, categorized into tertiles representing low, intermediate, and high genetic risk. Systemic metabolic biomarkers were quantified using high-throughput proton nuclear magnetic resonance (¹H-NMR) spectroscopy (Nightingale Health) ^19^; 249 metabolites underwent log(x+1) transformation and Z-score standardization.

Twelve quantitative cardiac phenotypes, encompassing atrial ^20^ and ventricular ^21^ structural remodelling, alongside electrophysiological indices ^22^, were extracted from concurrent cardiac magnetic resonance (CMR) (1.5T Siemens MAGNETOM Aera) and resting 12-lead electrocardiogram (ECG) (GE Cardiosoft system) data ^23^. Detailed definitions are provided in the **Supplementary Methods**.

### 2.4 Statistical Analysis

#### 2.4.1 Standard Epidemiological Modelling

All primary epidemiological analyses were performed across 10 multiply imputed datasets ^24^. Cox proportional hazards models estimated hazard ratios (HRs) for incident AF. Continuous fibrosis indices and liver cT1 were Z-standardized after 1^st^-99^th^ percentile winsorization. In the main cohort, Model 1 adjusted for age, sex, and ethnicity; Model 2 added BMI, smoking status, alcohol consumption frequency, and TDI; Model 3 further included clinical comorbidities; and Model 4 added the AF PRS and 10 genetic PCs. Imaging-based models additionally adjusted for PDFF.

#### 2.4.2 Metabolome-wide Causal Mediation Analysis

Within the NMR multi-omics subcohort (N = 362,050 with complete metabolomics), participants were partitioned into training (70%) and test (30%) sets. A two-step screening framework evaluated mediators: Path A (exposure-to-mediator) was estimated via linear regression adjusted for demographic and lifestyle covariates, and Path B (mediator-to-outcome) via Cox models with full clinical and genetic adjustments. Candidates meeting stringent dual thresholds for both FIB-4 and NFS (false discovery rate [FDR]-corrected *P* < 0.05, |β_Path A_| > 0.05 SD and HR_Path B_ outside the interval [0.95, 1.05]) formed a 144-metabolite candidate pool. Representative candidates underwent regression-based causal mediation analysis (*CMAverse* package ^25^), with average causal mediation effects (ACME) and standard errors estimated via the multivariate delta method ^26^.

#### 2.4.3 Feature Selection, Target-Organ Validation, and Phenotypic Clustering

Elastic Net framework ^27^ was implemented to the training set with 500 bootstraps; metabolites retained ≥ 75% of iterations were selected as the final core features (algorithm hyperparameters detailed in the **Supplementary Methods**).

Multivariable linear regressions were performed within the CMR and ECG subcohort—excluding pre-existing AF or severe cardiovascular disease prior to the imaging visit—to map the associations of the Elastic Net-derived features with the 12 quantitative cardiac phenotypes. These models were adjusted for age, as well as sex, systolic blood pressure, history of antihypertensive medication, and BMI.

Systemic metabolic phenotypes were derived using an unsupervised Gaussian mixture model (GMM; *mclust* package) ^28^ based on the final predictive features. The model was prespecified at three clusters with the covariance structure chosen by maximum Bayesian Information Criterion (BIC). Internal clustering stability was assessed via 500 bootstrap resamples using the adjusted Rand index (ARI). The final GMM was applied to the metabolomics subcohort to assign discrete phenotype probabilities, with the resulting clusters ordered chronologically by observed incident AF rates, and their geometric separation visualized via uniform manifold approximation and projection (UMAP) ^29^.

#### 2.4.4 Derivation and Validation of the Metabolomic Risk Score

A continuous MRS was constructed by fitting the final core features into a multivariable Cox proportional hazards model in the training cohort. Raw MRS was computed as the weighted linear combination of metabolites using unpenalized Cox regression coefficients, and then Z-score standardized. Joint effects, multiplicative interactions, and additive interactions (assessed via Relative Excess Risk due to Interaction [RERI] and Attributable Proportion) ^30^ between the MRS and the AF PRS were assessed in the full NMR cohort, with the independent test set used for out-of-sample confirmation. Incremental predictive value was evaluated by adding the MRS sequentially to established clinical models (CHARGE-AF ^31^, ARIC ^32^, and C2HEST ^33^), and combined clinical-genetic models. Predictive improvements across these stepwise models were quantified using time-dependent area under the curve (AUC) ^34^, calibration curves, continuous net reclassification improvement (cNRI), and integrated discrimination improvement (IDI) ^35^ at a 5-year follow-up horizon, with 95% confidence intervals and *P*-values derived via 500 bootstrap resamples.

#### 2.4.5 Subgroup and Sensitivity Analyses

Stratified analyses were conducted across prespecified subgroups defined by age, sex, BMI, diabetes status, alcohol consumption, MASLD, and metabolic dysfunction and alcohol-associated steatotic liver disease (MetALD) status. Sensitivity analyses included: (1) excluding AF cases within the first two years of follow-up; (2) excluding baseline cardiovascular disease, former/harmful drinkers, or haematological malignancies; (3) restricting to pure incident AF by the strict exclusion of atrial flutter; (4) applying alternative diagnostic liver fibrosis thresholds; (5) fitting Fine-Gray subdistribution hazard models ^36^ accounting for competing mortality risk; and (6) implementing inverse probability of treatment weighting (IPTW) ^37^ to balance baseline characteristics.

All analyses were performed using R software (version 4.5.1), with two-sided *P* < 0.05 considered statistically significant.

## 3. Results

### 3.1 Association of Hepatic Fibro-Inflammation with Incident AF

A total of 403,974 participants were included in the primary analysis, with 27,032 individuals in the liver-imaging subcohort **(Figure S1)**. Over a median follow-up of 13.18 years, 26,677 incident AF cases were identified in the primary cohort. Participants classified as high-risk for liver fibrosis were older, predominantly male, and had a higher cardiometabolic burden **(Tables S1-S5)**.

All biochemical liver fibrosis indices were associated with increased AF risk across all models. Individuals defined at high-risk for NFS and FIB-4 exhibited 54% (HR 1.54, 95% CI 1.43–1.66) and 53% (HR 1.53, 95% CI 1.44–1.62) elevated risk of AF, respectively. Clinically significant fibro-inflammation (cT1 ≥ 750 ms) conferred a 41% (HR 1.41, 95% CI 1.11–1.79) increased AF risk **(Table 1)**. Dose-response analyses **(Figure S2)** revealed nonlinear associations for all biochemical indices. AST/ALT showed a monotonic increase in AF risk, whereas APRI displayed a clear threshold effect. In contrast, liver cT1 exhibited a consistent dose-dependent relationship with incident AF.

**Table 1.**
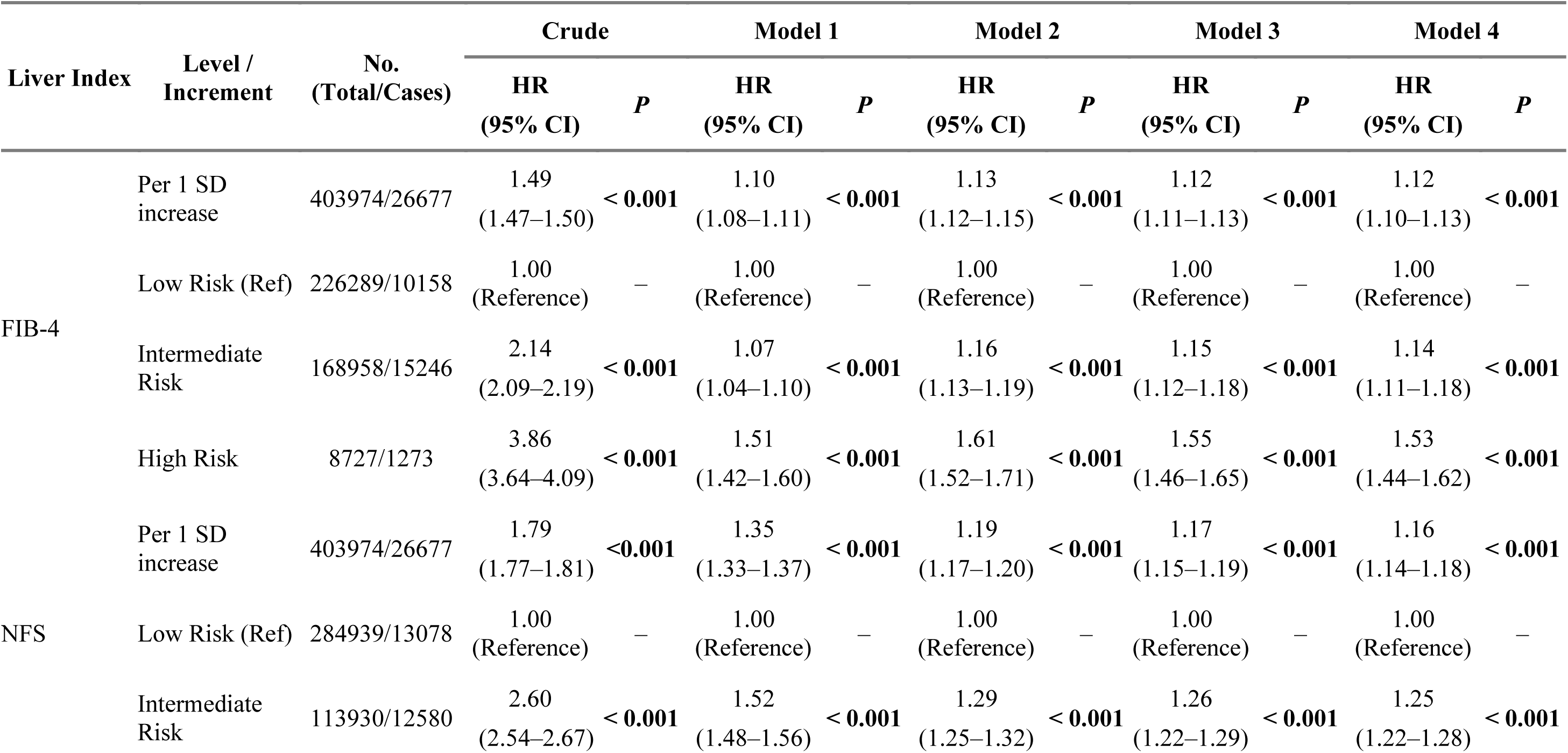
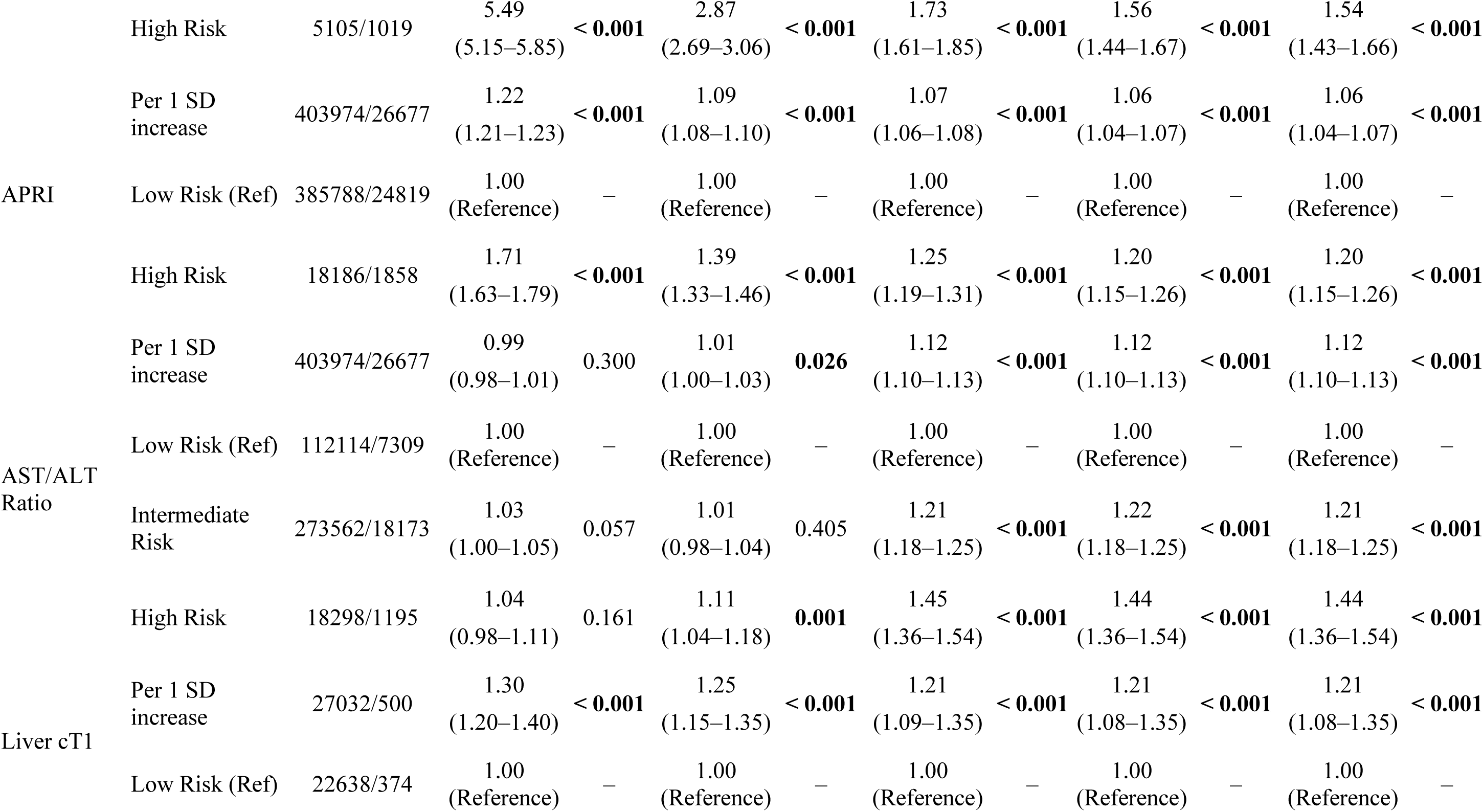
Stepwise multivariable associations between liver fibrosis indices and incident atrial fibrillation.

Subgroup analyses confirmed the robustness of these associations across clinically relevant strata (**Figures S3-S6**). Notably, associations were more pronounced among daily alcohol consumers and individuals without baseline diabetes or MASLD (all FDR-adjusted *P*_interaction_ < 0.05). Conversely, among participants with severe obesity (BMI ≥ 40 kg/m²), the predictive value of FIB-4 and APRI was attenuated and no longer statistically significant.

Sensitivity analyses further supported the stability of the findings **(Table S6)**. Associations remained consistent in a 2-year landmark analysis and after excluding participants with baseline cardiovascular disease, heavy alcohol consumption, or haematological malignancies. Results were also robust to alternative diagnostic thresholds for liver fibrosis indices and a stricter AF definition excluding atrial flutter. Finally, the observed associations persisted in competing risk models accounting for mortality and in IPTW analyses.

### 3.2 Metabolome-wide Causal Mediation Linking Hepatic Fibrosis to Incident AF

Given that circulating metabolites directly capture systemic inter-organ crosstalk, we identified 144 circulating metabolites linking hepatic fibrosis to incident AF **(Figure 1A)**. These metabolites showed both direct associations with AF and indirect mediation effects across liver fibrosis indices after multivariable adjustments.

**Figure 1.**
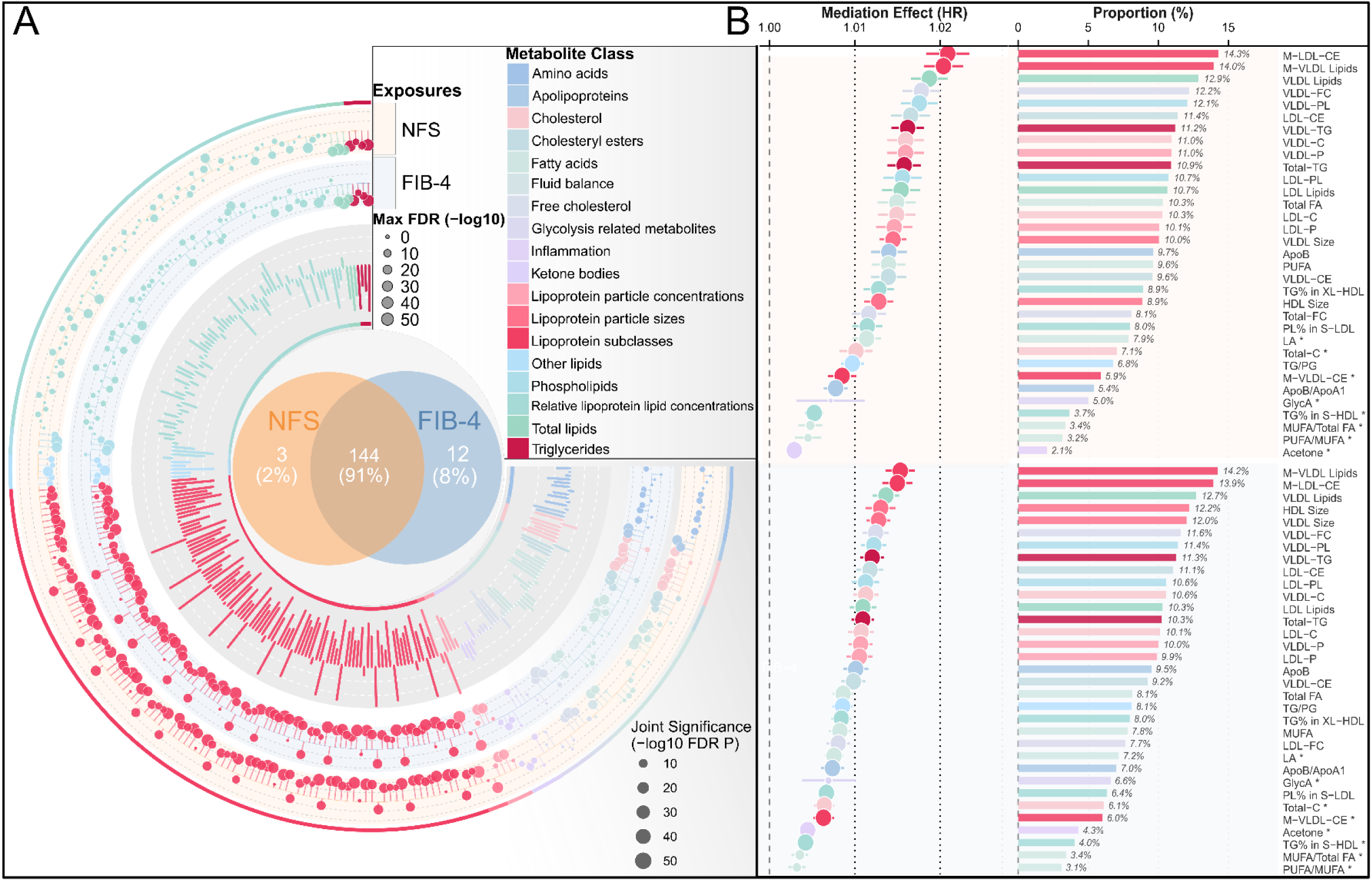
Metabolome-wide mediation atlas and identification of the core liver-heart axis metabolome. **(A)** Trivalent circular mediation atlas for 249 circulating metabolites. The two outer tracks visualize Path A effects (β) for NFS and FIB-4 (dashed lines: 0.25 SD increments); point sizes reflect the maximum FDR-adjusted *P*-value across Path A and Path B. The inner bar track represents Path B hazard ratios (HR) for incident AF (reference lines: HR 0.90-1.10). The central Venn diagram identifies 144 core mediators meeting significance criteria (dual FDR < 0.05, β_PathA_ > 0.05 SD, and HR thresholds), representing 91% of the union of significant mediators across both indices (percentages rounded; totals may differ by ±1%). **(B)** Mediation profiles of top-ranked core metabolites for NFS (top) and FIB-4 (bottom). Forest plots (left) and bar charts (right) illustrate the Average Causal Mediation Effect (ACME) and mediation proportions. Featured metabolites with ≥ 5% mediated proportion or classified as ultra-stable mediators. Point sizes indicate −log_10_ FDR *P*. *Note: During initial screening (Panel A), all models were performed in the training set (n = 253,434; 16,964 AF events). Path B models were adjusted for demographics, anthropometrics, lifestyle factors, comorbidities, lipid-lowering therapy, an AF-specific polygenic risk score (PRS), and 10 genetic principal components (PCs), whereas Path A models were adjusted for demographic and lifestyle covariates. Path A effects were estimated by linear regression and Path B effects by Cox proportional hazards regression. Formal causal mediation estimation (Panel B) was conducted in the full NMR subcohort (n = 362,050; 23,995 AF events), all clinical and genetic covariates were fully adjusted in both mediator and outcome models to satisfy sequential ignorability. Asterisks (*) denote ultra-stable mediators with a selection probability ≥ 75% across 500 bootstrap iterations using Elastic Net (α = 0.5).* **Abbreviations:** FIB-4, Fibrosis-4 index; NFS, NAFLD fibrosis score; FDR, false discovery rate; ACME, average causal mediation effect; VLDL-C/LDL-C, VLDL/LDL cholesterol; CE, cholesteryl esters; FC, free cholesterol; PL, phospholipids; TG, triglycerides; FA, fatty acids; MUFA/PUFA, monounsaturated/polyunsaturated fatty acids; LA, linoleic acid; GlycA, glycoprotein acetyls; LDL, low-density lipoprotein; VLDL, very-low-density lipoprotein; ApoA1, apolipoprotein A1; ApoB, apolipoprotein B; XS, very small; S, small; M, medium; L, large; XL, very large; XXL, extremely large particle-size prefixes. A circular mediation atlas of 249 plasma metabolites showing NFS and FIB-4 exposure effects and hazard ratios for incident atrial fibrillation in outer and inner tracks, with a central Venn diagram of 144 shared core mediators; adjacent forest and bar plots rank the top mediators by average causal mediation effect and mediation proportion.

Formal causal mediation analyses revealed that the relationships between chronic fibrosis/steatosis indices (FIB-4 and NFS) and incident AF were predominantly mediated by atherogenic lipoproteins and specific lipid subclasses (Figure 1B). Cholesteryl esters in medium low-density lipoprotein (LDL) and total lipids in medium very low-density lipoprotein (VLDL) were the strongest individual mediators, accounting for 13.9%–14.3% and 14.0%–14.2% of the total effect, respectively. Additionally, VLDL cholesterol, LDL cholesterol, and apolipoprotein B each mediated 9.5%–11.0% of the associations (**Figure 1B, Table S7**).

### 3.3 Machine Learning Prioritization and Target-Organ Validation

To distill the candidate pool into non-redundant core features, 8 core metabolites were prioritized from the 144 candidates via machine learning **(Figure S7)**. Mapping these metabolites to subclinical cardiac phenotypes revealed distinct morphological and electrophysiological correlations (**Figure 2A**). Specifically, elevated glycoprotein acetyls (GlycA), MUFA as a percentage of total fatty acids (MUFA%), and triglyceride percentage in small HDL (TG% in S-HDL) inversely correlated with cardiac chamber volumes and left ventricular mass, while positively correlated with resting heart rate. Linoleic acid demonstrated similar inverse associations with cardiac structure but minimal association with heart rate. Conversely, the polyunsaturated-to-monounsaturated fatty acid ratio (PUFA/MUFA) was positively correlated with chamber volumes and inversely associated with resting heart rate.

**Figure 2.**
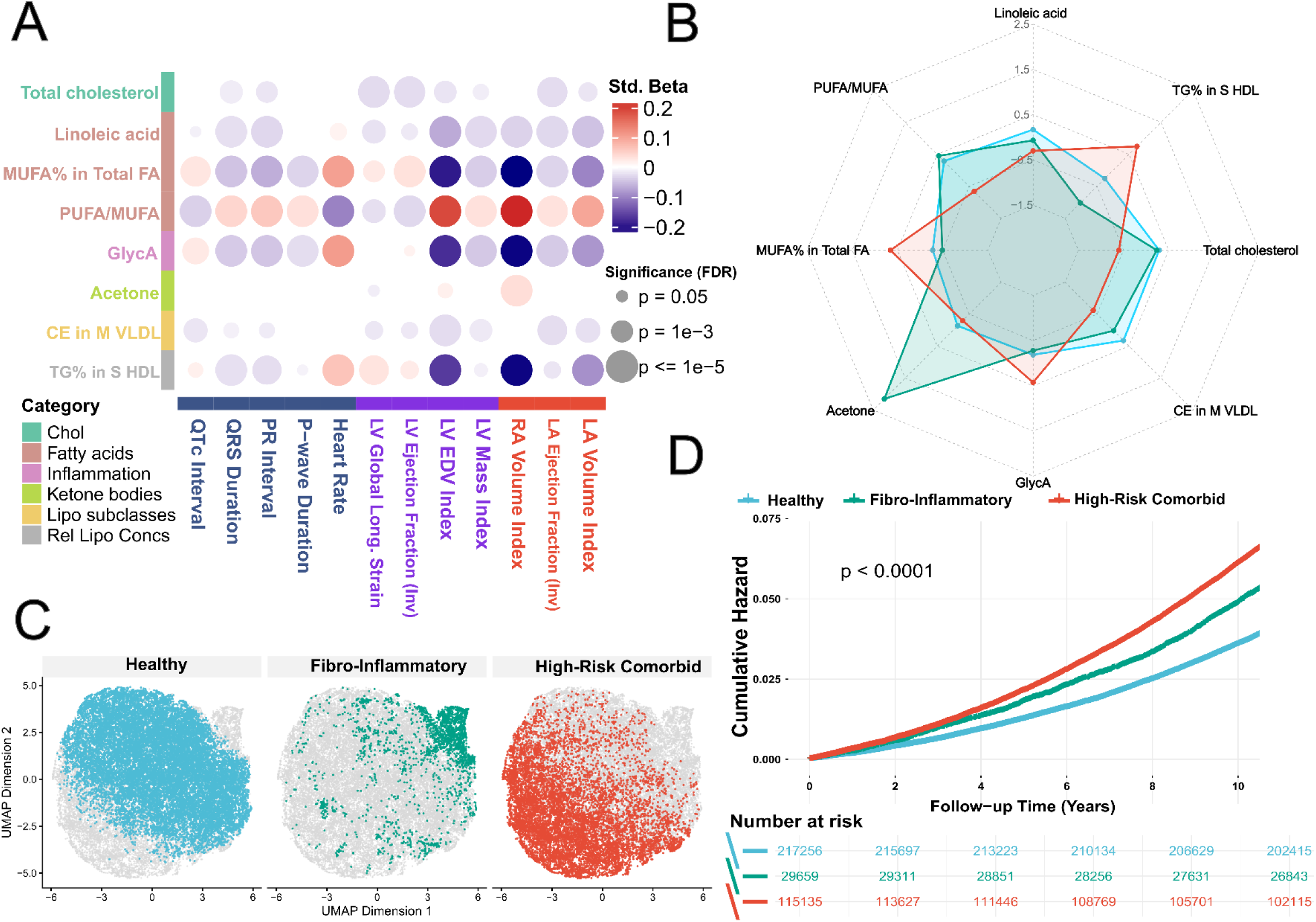
Cardiac pathophysiological profiling and clinical phenotyping of the ultra-stable core metabolites. **(A)** Associations between 8 core metabolites and cardiac structure/function. Bubble heatmap of multivariable linear regressions linking metabolites to cardiac magnetic resonance (CMR) and electrocardiogram (ECG) metrics at imaging visit (Cardiac Phenotype Subcohorts; n = 26,426 to 56,353 across the 12 cardiac metrics). Color gradient indicates standardized regression coefficient (β); point size denotes FDR-adjusted statistical significance. Models were adjusted for age, sex, systolic blood pressure, and antihypertensive medications, with additional BMI adjustment for non-BSA indexed targets. LVEF and LAEF were sign-inverted (-LVEF, -LAEF, denoted ‘Inv’) such that higher values correspond to worse function, ensuring a positive β indicates worsening function. **(B)** Metabolic profiles of Gaussian Mixture Model (GMM)-derived metabolic phenotypes. Radar chart displaying standardized mean abundance (Z-scores) of the 8 core metabolites across three clusters: Healthy (blue), Fibro-Inflammatory (green), and High-Risk Comorbid (red). **(C)** Uniform Manifold Approximation and Projection (UMAP) visualization of the three clusters, superimposed on a random background sample of the overall cohort (grey points). **(D)** Incident AF by metabolic phenotype. Kaplan-Meier curves demonstrating cumulative hazard of AF stratified by the three metabolic phenotypes (log-rank test). Following adjustment, the crude ordering inverts, with the Fibro-Inflammatory cluster showing the strongest independent association (Table S9). *Note: Clustering and survival analyses were performed in the NMR Multi-omics Cohort (n = 362,050; 23,995 incident AF events). KM curves are displayed to 10 years, the time at which all clusters retain ≥80% at-risk.* **Abbreviations:** AF, atrial fibrillation; PUFA, polyunsaturated fatty acids; MUFA, monounsaturated fatty acids; FA, fatty acids; GlycA, glycoprotein acetyls; CE, cholesteryl esters; TG, triglycerides; S HDL, small high-density lipoprotein; M VLDL, medium very low-density lipoprotein; Chol, cholesterol; Lipo subclasses, lipoprotein subclasses; Rel Lipo Concs, relative lipoprotein lipid concentrations; LV, left ventricular; LA, left atrial; RA, right atrial; EDV, end-diastolic volume; BMI, body mass index; BSA, body surface area; UMAP, Uniform Manifold Approximation and Projection; LVEF, left ventricular ejection fraction; LAEF, left atrial emptying fraction. Panel A, bubble heatmap of associations between 8 core metabolites and 12 cardiac MRI/ECG measures; Panel B, radar chart of metabolite Z-scores across three metabolic clusters (Healthy, Fibro-Inflammatory, High-Risk Comorbid); Panel C, UMAP projection of the three clusters; Panel D, Kaplan–Meier cumulative-hazard curves of incident AF by cluster with a number-at-risk table.

### 3.4 Unsupervised Clustering and Metabolic Stratification

To stratify underlying systemic metabolic phenotypes, unsupervised clustering based on the 8 core metabolites within the NMR multi-omics subcohort (N = 362,050) identified three distinct clusters: Healthy (N = 217,256), Fibro-Inflammatory (N = 29,659), and High-Risk Comorbid (N = 115,135), with a stable ARI of 0.717 (**Figure S8, Figure 2C**).

These phenotypes exhibited distinct metabolic distributions (**Figure 2B**). The Healthy phenotype was characterized by elevated levels of linoleic acid and cholesteryl esters in medium VLDL. The Fibro-Inflammatory phenotype was defined by elevated acetone and decreased TG% in S-HDL. Conversely, the High-Risk Comorbid phenotype exhibited increased MUFA%, GlycA, and TG% in S-HDL, alongside a relative depletion of the remaining core metabolites.

Despite comparable genetic susceptibility, age and overall comorbidity burden increased progressively from the Healthy to the High-Risk Comorbid phenotype; notably, the Fibro-Inflammatory cluster presented the lowest BMI and highest female proportion, yet the highest prevalence of high-risk fibrosis **(Table S8)**.

Compared with the Healthy group, both the Fibro-Inflammatory and High-Risk Comorbid groups exhibited higher cumulative risk of incident AF (as shown in **Figure 2D**, note reversed ordering post-adjustment) and remained significantly associated with incident AF after full adjustment (HR 1.19, 95% CI 1.14–1.25 and HR 1.04, 95% CI 1.01–1.07, respectively) **(Figure 2D, Table S9).** The High-Risk Comorbid cluster carried the highest unadjusted hazard (HR 1.65, 95% CI 1.61–1.70) but was largely attenuated after adjustment for adiposity, lifestyle, and comorbidities, whereas the Fibro-Inflammatory cluster was only modestly attenuated, reversing the crude ranking **(Table S9)**.

### 3.5 Derivation and Incremental Predictive Value of the Metabolomic Risk Score

To evaluate combined clinical-genetic utility, a continuous MRS was constructed based on the 8 core metabolites (**Figure 3A**). Among these, MUFA%, total cholesterol, and PUFA/MUFA carried the largest positive weights, whereas linoleic acid contributed a dominant inverse (protective) weight. Joint stratification by genetic (PRS) and metabolic (MRS) risk revealed a clear dose-response gradient (**Figure 3B, Table S10**). Using the low PRS/low MRS group as reference, the dual burden of high PRS and high MRS exhibited the greatest incident AF risk (HR 3.26, 95% CI 3.04–3.49). Notably, within the high PRS stratum, a low MRS attenuated AF risk (HR 2.75, 95% CI 2.56–2.96), indicating partial risk mitigation by favourable metabolic profiles. In the full cohort, metabolic and polygenic risk interacted to compound AF hazard on both the multiplicative (*P*_multiplicative_ < 0.001) and additive scales; elevated metabolic risk amplified genetic vulnerability in the intermediate-PRS (RERI 0.17, 95% CI 0.04–0.29) and high-PRS (RERI 0.17, 95% CI 0.01–0.33) strata. In the test cohort, this interaction was reproduced on the multiplicative scale (*P*_multiplicative_ = 0.026) but not the additive scale **(Table S11)**.

**Figure 3.**
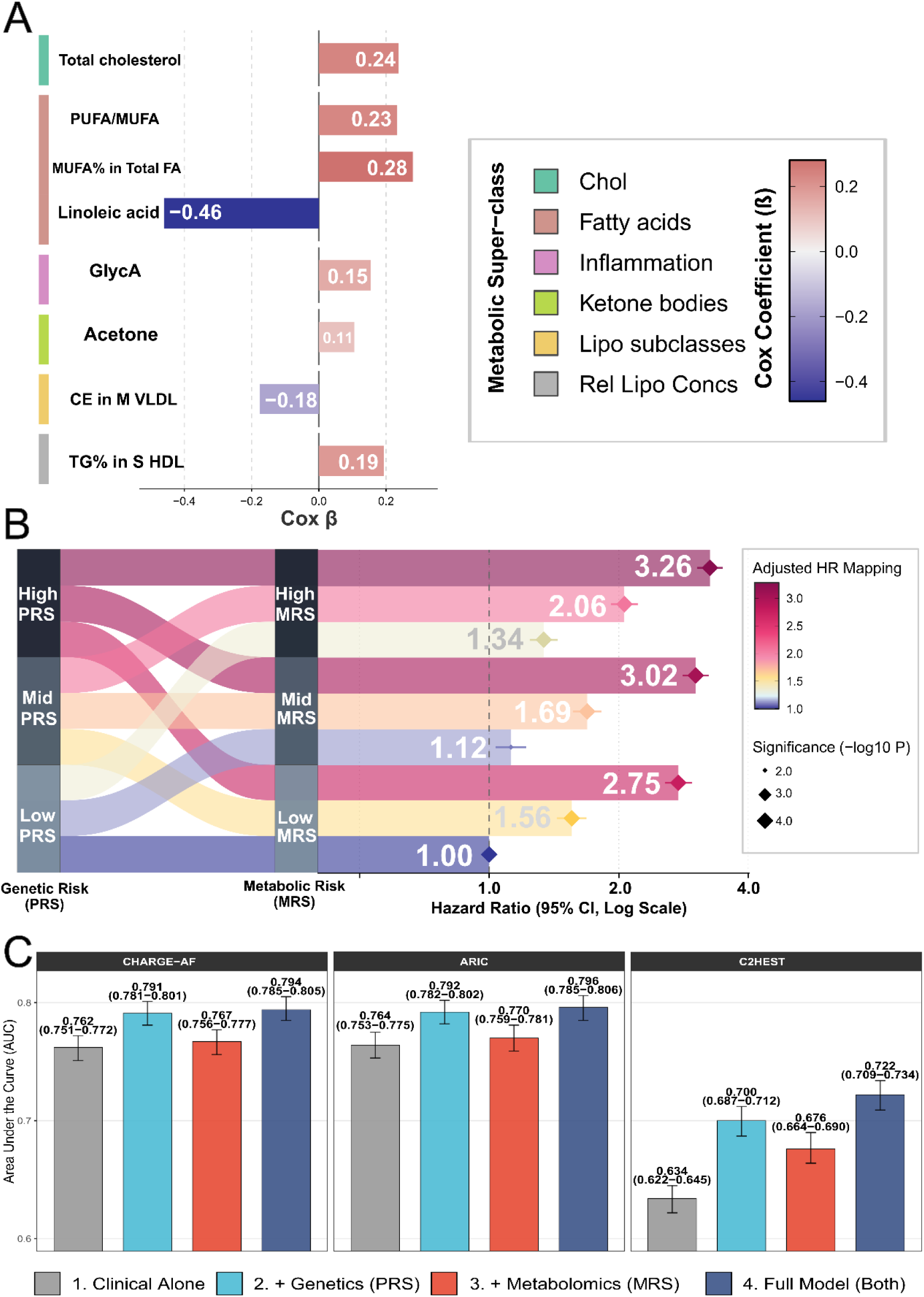
Derivation, joint risk stratification, and incremental predictive value of the Metabolomic Risk Score (MRS). **(A)** Derivation of the MRS. Unshrunken Cox coefficients (β) for the 8 core metabolites (training set; n = 253,434; 16,964 AF events). Colors indicate the effect magnitude and direction; the left vertical band denotes metabolic super-classes. **(B**) Joint genetic and metabolic risk for incident AF (NMR Multi-omics Cohort; n = 362,050; 23,995 incident AF events). Sankey diagram maps participant distribution across tertiles (Low, Mid, High) of the Polygenic Risk Score (PRS) and MRS. The accompanying forest plot shows adjusted hazard ratios (HR) and 95% confidence intervals (CI) for incident AF across the 9 joint categories, using Low PRS + Low MRS as reference. Point sizes reflect the statistical significance (−log10 P-value). **(C)** Incremental predictive performance (test cohort; n = 108,616; 7,031 incident AF events). Area Under the Curve (AUC) for 5-year AF prediction using CHARGE-AF, ARIC, and C2HEST models, sequentially updated with PRS, MRS, and the full model (PRS + MRS). Error bars represent 95% CIs derived from 500 bootstrap iterations. *Note: Models in Panel B were adjusted for demographics, anthropometrics, lifestyle factors, comorbidities, and the top 10 genetic principal components.* **Abbreviations:** AF, atrial fibrillation; HR, hazard ratio; CI, confidence interval; AUC, area under the curve; PUFA, polyunsaturated fatty acids; MUFA, monounsaturated fatty acids; FA, fatty acids; GlycA, glycoprotein acetyls; CE, cholesteryl esters; TG, triglycerides; M VLDL, medium very low-density lipoprotein; S HDL, small high-density lipoprotein; PRS, polygenic risk score; MRS, metabolomic risk score; CHARGE-AF, Cohorts for Heart and Aging Research in Genomic Epidemiology-Atrial Fibrillation; ARIC, Atherosclerosis Risk in Communities; C2HEST, coronary artery disease/chronic obstructive pulmonary disease (1 point each), hypertension, elderly (≥75 years), systolic heart failure, thyroid disease; Cox, Cox proportional hazards regression; Δ, change relative to baseline. Panel A, bar chart of Cox coefficients for the 8 metabolites in the metabolomic risk score; Panel B, Sankey diagram linking PRS and MRS tertiles with a forest plot of adjusted hazard ratios for the nine joint categories; Panel C, grouped bar chart of AUCs for CHARGE-AF, ARIC and C2HEST models updated with PRS, MRS and both.

Incorporation of the MRS into established clinical risk models improved discrimination while preserving adequate calibration (**Figure 3C**, **Figure 4A-F**). When added to the CHARGE-AF model, the MRS modestly improved discrimination (ΔAUC = +0.005; *P* < 0.002) and significantly enhanced risk reclassification (cNRI = 0.102, 95% CI 0.050–0.149; *P* < 0.001). These improvements persisted even after further adjustment for PRS (ΔAUC = +0.003; *P* < 0.002; cNRI = 0.093, 95% CI 0.041–0.139; *P* < 0.001) (**Figure 4G**, **Table S12**).

**Figure 4.**
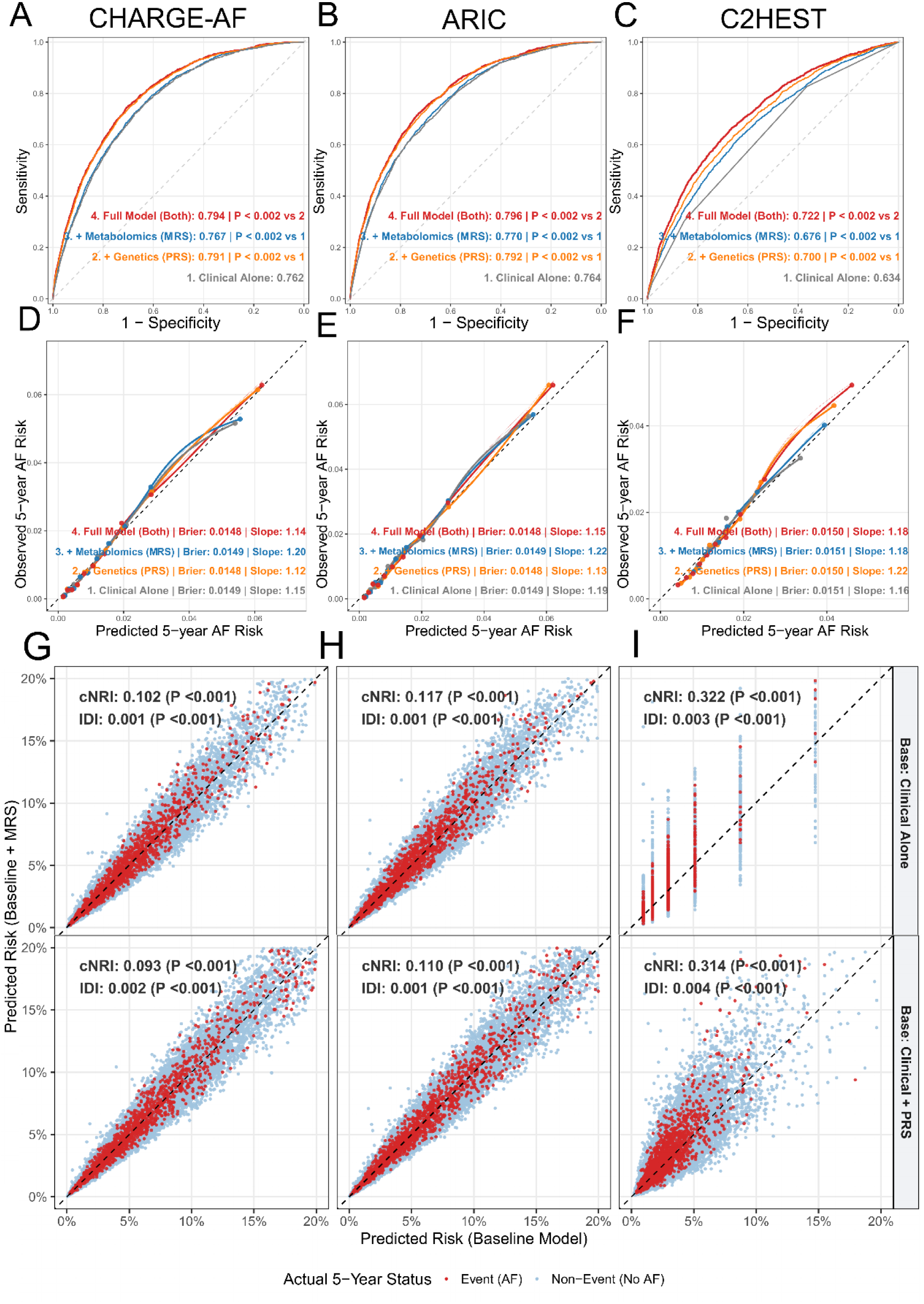
Incremental discrimination, calibration, and reclassification performance of the MRS for 5-year incident atrial fibrillation. **(A-C)** Time-dependent ROC curves evaluating 5-year AF prediction using CHARGE-AF (A), ARIC (B), and C2HEST (C). Baseline models (grey) were sequentially updated with the Polygenic Risk Score (PRS, orange), Metabolomic Risk Score (MRS, blue), and the full model combining both (red). Inset values indicate the Area Under the Curve (AUC) with bootstrap-derived *P*-values (500 iterations) for incremental improvement. **(D-F)** Calibration plots corresponding to (A-C), comparing predicted versus observed 5-year AF risk. Points represent risk deciles with loess smoothing curves. The dashed diagonal line indicates perfect calibration. Insets show the overall Brier score and calibration slope for each model. **(G-I)** Reclassification scatter plots demonstrating individual-level shifts in predicted risk after adding MRS to clinical models (top) and PRS to clinical baselines (bottom). Red dots represent individuals with incident AF (events), and blue dots represent those without AF (non-events). Points above the dashed diagonal line indicate increased predicted risk after adding the MRS. Insets report continuous Net Reclassification Improvement (cNRI) and Integrated Discrimination Improvement (IDI) metrics. *Note: All performance metrics were evaluated in the independent test cohort (n = 108,616; 7,031 incident AF events). P values report incremental improvement vs the indicated baseline model: ‘vs 1’ indicates comparison against the Clinical Alone baseline; ‘vs 2’ indicates comparison against the Clinical Alone + Genetics (PRS) model. A calibration slope > 1 indicates the model overestimates the variance of predicted risk relative to observed risk; ideal slope = 1. The column-like clustering of points in Panel I reflects the discrete, integer-valued nature of the baseline C2HEST score on the x-axis. ΔAUC and cNRI values represent the incremental predictive improvements compared to the respective baseline clinical-alone models. Brier score, mean squared difference between predicted probabilities and observed outcomes; slope, calibration slope; Δ, change relative to baseline.* **Abbreviations:** ROC, Receiver operating characteristic; AF, atrial fibrillation; PRS, polygenic risk score; MRS, metabolomic risk score; CHARGE-AF, Cohorts for Heart and Aging Research in Genomic Epidemiology-Atrial Fibrillation; ARIC, Atherosclerosis Risk in Communities; C2HEST, coronary artery disease/chronic obstructive pulmonary disease (1 point each), hypertension, elderly (≥75 years), systolic heart failure, thyroid disease; IDI, integrated discrimination improvement; cNRI, continuous net reclassification improvement. Panels A–C, time-dependent ROC curves for 5-year AF prediction with CHARGE-AF, ARIC and C2HEST baselines updated by PRS, MRS and the full model; Panels D–F, corresponding calibration plots of predicted versus observed risk; Panels G–I, reclassification scatter plots of predicted risk before and after adding MRS or PRS, with cNRI and IDI insets.

Comparable improvements were observed with the ARIC and C2HEST models (**Figure 4H-I, Table S12**). Notably, incorporation of the MRS into the C2HEST model yielded a larger gain in discrimination and reclassification (ΔAUC = +0.042; *P* < 0.002; cNRI = 0.322, 95% CI 0.272–0.367; *P* < 0.001), with similar enhancements observed for the ARIC model.

## 4. Discussion

### 4.1 Principal Findings

In this large-scale prospective study, we delineate the trajectory from hepatic fibro-inflammation to incident AF. First, advanced liver fibro-inflammation, assessed by biochemical indices and cT1, was independently associated with AF. Second, the liver-heart axis operates via a dual-track mechanism: circulating lipoproteins account for much of the indirect effect, while eight prioritized “hub” metabolites map onto distinct electrophysiological and structural cardiac signatures. Third, we identify a high-risk “Fibro-Inflammatory” phenotype characterized by a lean and normolipidemic profile but adverse metabolic perturbations. Fourth, the derived MRS demonstrated a “double-hit” interaction with PRS. Finally, the MRS provides meaningful reclassification beyond established clinical risk scores, with the greatest benefit observed in parsimonious models.

### 4.2 Hepatic Fibro-Inflammation as an Independent Contributor to AF

These findings extend prior liver-heart axis observations linking MASLD with AF risk ^6^, by distinguishing fibro-inflammation from simple steatosis. Using cT1 mapping, clinically significant fibro-inflammation was associated with a 41% higher AF risk independent of liver fat content assessed by PDFF, supporting the emerging view that fibrotic and inflammatory components—rather than lipid accumulation alone—drive cardiovascular risk ^38^. Subgroup analyses further revealed effect heterogeneity. Associations for FIB-4 and APRI were strongest among individuals without diabetes or MASLD but attenuated in severe obesity. This divergence highlights competing pathophysiological mechanisms: in metabolically healthier individuals, early hepatic fibro-inflammation may act as a relatively unconfounded arrhythmogenic driver, whereas in severe obesity, structural and haemodynamic factors—including atrial stretch and epicardial adiposity ^39, 40^—may dominate.

### 4.3 Dual-Track Liver-Heart Axis: Molecular Mediators and Target-Organ Effects

At the molecular level, a “quantitative versus qualitative” paradox was observed. While VLDL- and LDL-derived lipids contributed the largest mediation effects, reflecting the atherosclerotic pathways linked to hepatic disease ^38^, these circulating lipids are highly collinear. Penalized selection reduced 144 candidates to eight non-redundant metabolites, prioritizing distinct signals over shared variance, consistent with established metabolomic frameworks ^8^. These metabolites capture complementary aspects of hepatic metabolic stress, including systemic inflammation (GlycA), lipid remodelling (TG% in S-HDL), and ketogenic stress (acetone). Importantly, mapping to cardiac phenotypes revealed a dual-track model diverging from the traditional structural paradigm ^41^.

The electrophysiological track, indexed by elevated GlycA, MUFA%, and TG% in S-HDL, was associated with higher resting heart rate, prolonged corrected QT (QTc), shortened PR/QRS intervals, and reduced chamber volumes. This profile aligns with inflammation- and lipotoxicity-related ion-channel and conduction abnormalities described in experimental models ^42^, though causality remains unconfirmed. Such “non-structural” mechanisms have also been proposed in metabolic AF and epicardial adiposity pathways ^43^.

In contrast, the structural track, characterized by elevated PUFA/MUFA ratio and depleted linoleic acid, was associated with atrial and ventricular chamber enlargement. Linoleic acid showed consistent inverse associations with cardiac volumes, supporting a cardioprotective role ^44^. The PUFA/MUFA signal likely reflects MUFA exhaustion superimposed on linoleic acid depletion, rather than a true PUFA surge, potentially influencing membrane dynamics and fibrotic remodelling in preclinical studies ^45^, though human validation is pending. Together, these findings associate hepatic dysfunction with cardiac changes, with electrical and structural tracks appearing dissociated.

### 4.4 A High-Risk Fibro-Inflammatory Phenotype Not Captured by Conventional Frameworks

Clustering analysis identified a clinically under-recognized high-risk phenotype. The Fibro-Inflammatory cluster—despite a lower BMI and minimal dyslipidemia—harbored the highest burden of advanced hepatic fibrosis (marked by an elevated AST/ALT ratio and lower platelets; **Table S8)** and carried a largely unattenuated AF risk (HR 1.19, 95% CI 1.14–1.25). In contrast, while the High-Risk Comorbid cluster was characterized by elevated GlycA, MUFA%, and TG% in S-HDL, its crude excess risk was largely accompanied by adiposity and comorbidities and substantially attenuated after adjustment, in keeping with a conventional metabolic syndrome–associated phenotype. This Fibro-Inflammatory cluster reflects hepatic energy decompensation rather than caloric excess, characterized by elevated acetone and reduced TG% in S-HDL. Progressive fibrosis disrupts ketogenesis ^46^, prompting compensatory extrahepatic ketone utilization alongside impaired lipoprotein regulation ^47^. Although acetone lacked pronounced correlations in the CMR-ECG phenomapping analysis within the relatively healthy subcohort, ketones may support myocardial energetics but also alter mitochondrial function in stressed myocardium ^48^, with potential electrophysiology implications yet to be verified within the Fibro-Inflammatory cluster.

These findings parallel lean MASLD, which carries a cardiovascular risk comparable to or exceeding that of typical MASLD despite favourable anthropometry ^49^. Notably, this phenotype (low BMI, normal lipids, and absent cardiometabolic comorbidity) is poorly captured by legacy AF risk scores (CHARGE-AF, ARIC, and C2HEST), which rely heavily on BMI, blood pressure, and overt cardiometabolic disease, consistent with our reclassification results.

### 4.5 Genome-Metabolome Interaction and Incremental Clinical Value

Integration of PRS and MRS highlights the interplay between inherited and acquired risk. Although AF is strongly heritable ^18^, the addition of MRS revealed a synergistic “double-hit” effect, with significant multiplicative interaction (*P*_multiplicative_ < 0.001) and additive interaction across the intermediate-to-high genetic spectrum; the multiplicative interaction was reproduced in the independent test cohort.

This suggests that hepato-metabolic dysfunction compounds underlying genetic susceptibility across the entire polygenic spectrum. These observations support combined genome–metabolome risk stratification as a more comprehensive framework for AF ^18, 50^.

From a translational perspective, MRS improved prediction across established clinical models, with a discrimination–reclassification dissociation. In CHARGE-AF and ARIC, discrimination gains were modest but reclassification was meaningful (continuous NRI ≈ 10–12%), consistent with AUC saturation effects ^35^. In contrast, the parsimonious C2HEST model showed substantially greater improvement (cNRI 32.2%), reflecting greater residual variance available for molecular augmentation.

Importantly, MRS retained incremental value even when added to PRS-enhanced models across all three scores, supporting the metabolome as an independent and modifiable risk axis distinct from both clinical and genetic domains ^8^. Clinical, polygenic, and metabolomic scores thus capture distinct, complementary axes of AF risk—current phenotype, inherited liability, and the current (and potentially modifiable) metabolic state.

### 4.6 Limitations

Several limitations merit consideration. First, the observational design precludes causal inference despite adjustment for polygenic risk; residual confounding and reverse causation remain possible. Second, the predominantly European ancestry of UK Biobank cohort limits generalizability across East Asian and African-ancestry populations presenting with different baseline lipid distributions and AF epidemiology. Third, the NMR platform does not capture the full metabolomic landscape, and additional omics layers may yield further insights ^19^. Finally, metabolomic profiling was cross-sectional; longitudinal multi-omic resampling is needed to assess dynamic trajectories and therapeutic responsiveness of the MRS.

## 5. Conclusion

Advanced hepatic fibro-inflammation is associated with incident AF through a dual-track mechanism separating electrophysiological from structural remodelling. The derived metabolomic risk score compounds polygenic risk and improves reclassification beyond established clinical tools, supporting the metabolome as an independent and potentially modifiable axis for AF risk stratification.

## CRediT Author Statement

**Guanyu Lin**: Investigation, Data curation, Formal analysis, Writing - original draft. **Jinzhu Hu**: Formal analysis (interpretation of data), Writing - review & editing. **Tongsheng Huang**: Formal analysis (interpretation of data), Writing - review & editing. **Wenli Gu**: Formal analysis (interpretation of data), Writing - review & editing. **Jingfeng Wang**: Formal analysis (interpretation of data), Writing - review & editing. **Yiyang Cao**: Formal analysis (interpretation of data), Writing - review & editing. **Linghua Fu**: Formal analysis (interpretation of data), Writing - review & editing. **Zhaoyu Liu**: Formal analysis (interpretation of data), Writing - review & editing. **Weiwen Lim**: Formal analysis (interpretation of data), Writing - review & editing. **Ching Chi-Keong**: Formal analysis (interpretation of data), Writing - review & editing. **Chrishan Ramachandra**: Formal analysis (interpretation of data), Writing - review & editing. **Hualin Fan**: Formal analysis (interpretation of data), Writing - review & editing. **Yinhua Zhang**: Formal analysis (interpretation of data), Writing - review & editing. **Shuai Wei**: Formal analysis (interpretation of data), Writing - review & editing. **Haifeng Zhang**: Formal analysis (interpretation of data), Writing - review & editing. **Yuan Jiang**: Formal analysis (interpretation of data), Writing - review & editing. **Yuling Zhang**: Formal analysis (interpretation of data), Writing - review & editing. **Lin Zhang**: Formal analysis (interpretation of data), Writing - review & editing. **Wengen Zhu**: Formal analysis (interpretation of data), Writing - review & editing. **Peng Yu**: Formal analysis (interpretation of data), Writing - review & editing. **Xiao Liu**: Conceptualization, Funding acquisition, Supervision, Project administration, Writing - review & editing. **Yangxin Chen**: Formal analysis (interpretation of data), Supervision, Writing - review & editing. **Derek John Hausenloy**: Formal analysis (interpretation of data), Supervision, Writing - review & editing.

## Acknowledgements

We extend our sincere gratitude to all the participants of the UK Biobank for their invaluable contribution to this resource. We also thank the UK Biobank staff for their dedication to establishing and maintaining this database. During the preparation of this work, the authors used Gemini and Claude for manuscript language polishing. After using these tools, the authors reviewed and edited the content as needed and took full responsibility for the content of the published article.

## Data availability statement

This research was conducted using the UK Biobank Resource under Application No. 366548. The data underlying this article are available from the UK Biobank upon approval of a research application.

## Supplemental Material

Table S1-S13 & Figure S1-S8

