## Supplementary Appendix (Tables S1-S13, Figures S1-S8) for "Hepatic Fibro-Inflammation and Atrial Fibrillation: A Dual-Track Metabolic Axis Revealed by a Metabolomic Clock"

**TABLE OF CONTENTS**

**SUPPLEMENTARY METHODS**

**SUPPLEMENTARY TABLES**

**SUPPLEMENTARY FIGURES**

#### Section 1. Key Variables Definitions and Clinical Outcome Ascertainment

Baseline clinical comorbidities were ascertained through a comprehensive, multi-source integration strategy to minimize misclassification. We integrated self-reported medical conditions (UK Biobank verbal interview, Field 20002), self-reported medication usage at Instance 0, biochemical/physical measurements, and linked hospital inpatient records encompassing both ICD-9 and ICD-10 coding systems. The precise diagnostic criteria utilized for covariates and outcomes were defined as follows:

- **Prevalent and Incident Atrial Fibrillation (AF):** Incident AF (the primary outcome) was ascertained via linked hospital inpatient records and death registries using ICD-10 code I48 and ICD-9 code 427.3. Participants with prevalent AF at baseline—defined rigorously by any occurrence of ICD-10 (I48) or ICD-9 (427.3) codes prior to or on the date of baseline assessment, or self-reported AF/atrial flutter—were strictly excluded from the primary longitudinal analysis.
- **Hypertension:** Defined by (1) hospital records (ICD-10: I10–I15; ICD-9: 401–405); (2) self-reported hypertension; (3) baseline use of blood pressure-lowering medications (e.g., beta-blockers, calcium channel blockers, ACE inhibitors, ARBs, or diuretics); or (4) automated blood pressure readings demonstrating a mean systolic blood pressure $\geq$ 140 mmHg or diastolic blood pressure $\geq$ 90 mmHg.
- **Dyslipidemia:** Defined by (1) hospital records (ICD-10: E78; ICD-9: 272); (2) self-reported high cholesterol; (3) baseline use of lipid-lowering therapies (e.g., statins, fibrates, ezetimibe); or (4) biochemical evidence at baseline (Triglycerides $\geq$ 1.7 mmol/L, or HDL cholesterol $<$ 1.03 mmol/L for males and $<$ 1.29 mmol/L for females).
- **Type 2 Diabetes Mellitus (T2DM):** To ensure high specificity for T2DM, we implemented a strict exclusion-based diagnostic pipeline. Participants with evidence of Type 1 diabetes or gestational diabetes (ICD-10: E10, O24.4, O24.8, O24.9, 648.8; ICD-9: 250.1, 250.3, 250.5, 250.7, 250.9; or self-report) were excluded from the T2DM cohort. T2DM was confirmed by any of the following: (1) hospital records (ICD-10: E11; ICD-9: 250.0, 250.2, 250.4, 250.6, 250.8); (2) self-reported Type 2 diabetes; (3) baseline use of glucose-lowering medications; or (4) biochemical thresholds including Glycated haemoglobin (HbA1c) $\geq$ 48 mmol/mol or random glucose $\geq$ 11.1 mmol/L.
- **Chronic Kidney Disease (CKD):** Defined by (1) hospital records (ICD-10: N18; ICD-9: 585); (2) self-reported renal failure; or (3) an estimated glomerular filtration rate (eGFR) $<$ 60 mL/min/1.73 $m^{2}$. The eGFR was calculated using the standardized CKD-EPI (Chronic Kidney Disease Epidemiology Collaboration) 2009 creatinine equation based on baseline serum creatinine levels.
- **Prevalent Cardiovascular Disease (CVD):** Defined broadly via hospital records or self-reports encompassing coronary heart disease and myocardial infarction (ICD-10: I20–I25; ICD-9: 410–414; self-report), heart failure (ICD-10: I50; ICD-9: 428; self-report), valvular heart disease (ICD-10: I05–I08, I34–I39; ICD-9: 394–396, 424; self-report), peripheral vascular disease (ICD-10: I70–I73; ICD-9: 440–443; self-report), and stroke (ICD-10: I60–I64; ICD-9: 430, 431, 433, 434, 436; self-report).

- **Metabolic Dysfunction-Associated Steatotic Liver Disease (MASLD):** MASLD was defined as the presence of hepatic steatosis in the absence of viral hepatitis. In the primary cohort (baseline), hepatic steatosis was identified through established hospital diagnostic records (ICD-10: K76.0, K75.8; ICD-9: 571.8) or a high-risk Fatty Liver Index (FLI) score $\geq$ 60. The FLI was calculated using baseline metabolic parameters: $FLI=\left( e^{L} \right)/\left( 1+e^{L} \right)\times100$, where $L=0.953\times\ln\left( Triglycerides \left[ mg/dL \right] \right)+0.139\times BMI \left[ kg/m^{2} \right]+0.718\times\ln\left( GGT \left[ U/L \right] \right)+0.053\times Waist circumference \left[ cm \right]-15.745$. For the liver imaging subcohort (Instance 2), to ensure strict temporal alignment, MASLD was dynamically redefined by the gold-standard imaging threshold of hepatic proton density fat fraction (PDFF) $\geq$ 5%, concurrently excluding any historical or newly incident viral hepatitis up to the date of the MRI scan.
- **Weekly Alcohol Intake:** Weekly alcohol consumption (grams/week) across respective visits was quantified by multiplying self-reported intake frequencies of specific alcoholic beverages by standard UK Biobank alcohol unit conversions (red and white wine: 12 g/glass; beer/cider: 16 g/pint; spirits, fortified wine, and other beverages: 8 g/measure).
- **metabolic dysfunction and alcohol-associated steatotic liver disease (MetALD):** MetALD was defined as participants meeting the respective MASLD criteria who concurrently consumed greater amounts of alcohol. In the primary cohort, this was strictly defined as 140–350 g/week for females and 210–420 g/week for males at baseline. In the imaging subcohort, this was dynamically re-evaluated using Instance 2 data, requiring either the same sex-specific weekly thresholds or a synchronous self-reported "Daily or almost daily" intake frequency.
- **Former and Harmful Drinking (For Sensitivity Analyses):** To eliminate potential confounding from direct cardiotoxic and hepatotoxic effects in our sensitivity analyses, participants with extreme alcohol consumption behaviors were systematically excluded. Specifically, "harmful drinkers" were defined according to established high-risk thresholds: alcohol consumption > 280 g/week for females and > 400 g/week for males. "Former drinkers" were strictly classified based on the self-reported "Previous" status in the UK Biobank baseline alcohol drinking status questionnaire (Field 1558). Furthermore, to mitigate reverse causality, a 2-year landmark analysis was performed, comprehensively excluding any participants who developed incident atrial fibrillation within the first two years of follow-up.

The requisite biochemical and hematological parameters were quantified using Beckman Coulter AU5800 and DxH 800 auto-analyzers, respectively, under rigorous central quality control (coefficients of variation consistently <5%) ^1, 2^. The biochemical indices for hepatic fibro-inflammation were calculated using baseline serum biomarkers according to the following established clinical formulas:

- **Fibrosis-4 (FIB-4) index ^3^=** $\left( \text{Age}\boldsymbol{\times}\text{AST} \right)\boldsymbol{/}\left( \text{Platelet}\boldsymbol{\times}\sqrt{\text{ALT}} \right)$
- **NAFLD Fibrosis Score (NFS) ^4^=**$\boldsymbol{-1.675+0.037}\boldsymbol{\times}\boldsymbol{Age}\left[ \boldsymbol{years} \right]\boldsymbol{+0.094}\boldsymbol{\times}\boldsymbol{BMI}\left[ \boldsymbol{kg}\boldsymbol{/}\boldsymbol{m}^{\boldsymbol{2}} \right]\boldsymbol{+1.13}\boldsymbol{\times}\boldsymbol{T}\boldsymbol{2}\boldsymbol{DM}\left[ \boldsymbol{yes=1,no=0} \right]\boldsymbol{+0.99}\boldsymbol{\times}\left( \boldsymbol{AST}\boldsymbol{/}\boldsymbol{ALT ratio} \right)\boldsymbol{-0.013}\boldsymbol{\times}\boldsymbol{Platelet Count}\left[ \boldsymbol{1}\boldsymbol{0}^{\boldsymbol{9}}\boldsymbol{/}\boldsymbol{L} \right]\boldsymbol{-0.66}\boldsymbol{\times}\boldsymbol{Albumin}\left[ \boldsymbol{g}\boldsymbol{/}\boldsymbol{dL} \right]$
- **AST to Platelet Ratio Index (APRI) ^5^=** $\left[ \left( \text{AST}\boldsymbol{/}\text{ULN} \right)\boldsymbol{/}\text{Platelet} \right]\boldsymbol{\times}\boldsymbol{100}$ **where ULN represents the upper limit of normal for AST (defined as 40 U/L in this study).**
- **AST/ALT ratio (De Ritis ratio) ^6^=** $\text{AST}\boldsymbol{/}\text{ALT}$

To comprehensively evaluate subclinical target-organ damage along the liver-heart axis, 12 quantitative cardiac phenotypes were extracted from the imaging subcohort (Instance 2). These phenotypes were categorized into three distinct pathophysiological domains:

- **Atrial Remodeling:** Evaluated using the left atrial volume index (LAVI), right atrial volume index (RAVI), and left atrial ejection fraction (LAEF).
- **Ventricular Remodeling:** Assessed via the left ventricular (LV) mass index, LV end-diastolic volume index (LVEDVI), LV ejection fraction (LVEF), and global longitudinal strain (GLS).
- **Electrophysiological Indices:** Extracted from concurrent resting 12-lead electrocardiograms, including resting heart rate, P-wave duration, PR interval, QRS duration, and corrected QT (QTc) interval.

*(Note: Parameters dependent on body size were strictly indexed to body surface area [BSA].)*

#### Section 2. Genetic Data Processing and Polygenic Risk Score (PRS) Construction

Genetic predisposition to AF was assessed using the pre-calculated, standard polygenic risk score (PRS) provided by the UK Biobank (Data-Field 26213). This standardized PRS was centrally derived by the UK Biobank and Genomics plc using a strictly quality-controlled pipeline. The scoring algorithm leveraged summary statistics from a large-scale genome-wide association study (GWAS) meta-analysis for AF ^7^. The raw PRS values were subsequently Z-score standardized across the analytical cohort. Participants were then stratified into low, intermediate, and high genetic risk categories based on the tertiles of this standardized score.

### **Section 3. Nuclear Magnetic Resonance (**${}^{\mathbf{1}}$**H-NMR) Metabolomics Acquisition and Preprocessing**

Systemic metabolic biomarkers (UK Biobank Category 220) were quantified from randomly selected EDTA plasma samples (aliquot 3) using a high-throughput proton nuclear magnetic resonance (${}^{1}$H-NMR) biomarker profiling platform (Nightingale Health Ltd., Finland) ^8^. The quantified biomarkers comprehensively span multiple systemic metabolic pathways, encompassing absolute concentrations and ratios of lipoprotein lipids distributed across 14 subclasses, various fatty acids, and low-molecular-weight metabolites including amino acids, ketone bodies, and glycolysis intermediates. Data acquisition spanned three phases from June 2019 to November 2023. Crucially, to ensure rigorous measurement consistency, our study utilized the updated July 2023 UK Biobank data release, which incorporates essential spectrometer recalibrations for the Phase 1 cohort ^9, 10^.

Regarding statistical preprocessing, due to the inherent right-skewness of most metabolic distributions, all continuous variables were subjected to a $\log\left( x+1 \right)$ transformation. Subsequently, Z-score standardization (mean = 0, standard deviation = 1) was applied to uniformly scale the features. This strict pre-processing protocol was executed to facilitate robust effect size computations, penalty applications in Elastic Net regression ^11^, and reliable geometric distance calculations in downstream unsupervised clustering models ^12^.

#### Section 4. Machine Learning Hyperparameters

To ensure full reproducibility of the analytical pipeline, the algorithm-specific hyperparameters and settings excluded from the main text are detailed as follows: The feature selection was conducted using a Cox proportional hazards Elastic Net framework (family = "cox") to rigorously account for the time-to-event nature of incident AF. Analyses were performed using the glmnet R package (version 4.1.10). The penalty mixing parameter ($\alpha$) was set to 0.5, and the convergence threshold was maintained at the package default of $1\times{10}^{-7}$. The 500 bootstrap resamples were initialized with a fixed random seed (seed = 2026). For each iteration, the optimal penalization parameter ($\lambda$) was selected using the strict 1-standard-error rule (lambda.1se) via 10-fold cross-validation.

#### Section 5. Calculation of Established Clinical Baseline Risk Scores

To rigorously evaluate the incremental prognostic value of the derived Metabolomic Risk Score (MRS), three established clinical algorithms for AF prediction were calculated for each participant based on baseline characteristics. To prevent exponential inflation and precisely estimate absolute risk, the continuous covariates in the CHARGE-AF and ARIC models were strictly mean-centered according to their original derivation cohorts. The specific mathematical formulations employed were as follows:

- **CHARGE-AF Score ^13^:** To calculate the 5-year absolute risk of incident AF, the linear predictor ($\Sigma\beta X$) was first computed using the following formula: $\Sigma\beta X=0.508\times\frac{Age-57}{5}+0.248\times\text{White race}+0.115\times\frac{Height-167}{10}+0.197\times\frac{Weight-79}{15}+0.458\times\frac{SBP-120}{20}-0.352\times\frac{DBP-75}{10}+0.359\times\text{Current smoker}+0.349\times\text{Antihypertensive medication}+0.237\times\text{Diabetes}+0.496\times\text{Myocardial infarction}+1.212\times\text{Heart failure}$
- **ARIC Score ^14^:** The 10-year absolute risk was estimated using the formula: $Risk_{10y}=1-{0.929}^{\exp\left( \Sigma\beta X \right)}$. To faithfully adapt the original ARIC derivation model to the UK Biobank demographic structure, the linear predictor ($\Sigma\beta X$) was calculated using population-centered variables: $\Sigma\beta X = 0.106 \times\left( Age-54.2 \right) - 0.191 \times\left( White race-0.77 \right) + 0.057 \times\left( Height-168.1 \right) + 0.418 \times\left( Current smoker-0.26 \right) + 0.013 \times\left( SBP-122.5 \right) + 0.428 \times\left( Antihypertensive medication-0.23 \right) + 0.285 \times\left( Diabetes-0.08 \right) + 0.551 \times\left( Coronary artery disease-0.05 \right) + 1.054 \times\left( Heart failure-0.01 \right)$

*(Note: Due to the unavailability of routine echocardiography and phonocardiography at the UK Biobank baseline, left ventricular hypertrophy and cardiac murmurs from the original ARIC score were replaced by the highly correlated diagnosis of coronary artery disease to maintain the model's overall prognostic integrity).*

- **C2HEST Score ^15^:** A simplified clinical point-based scoring system calculated as the arithmetic sum of the following clinical features: Coronary Artery Disease (1 point), Chronic Obstructive Pulmonary Disease (1 point), Hypertension (1 point), Elderly (Age $\geq$ 75 years, 2 points), Heart Failure (2 points), and Thyroid Disease (1 point). Total scores ranged from 0 to 8 points.

#### Section 6. Statistical Framework for Competing Risk and Causal Inference

To rigorously evaluate whether the observed pro-arrhythmic effect of liver injury was confounded by survival bias, we employed Fine-Gray subdistribution hazard models ^16^. In these models, all-cause mortality prior to incident AF was treated as a competing event. Furthermore, to closely mimic the exchangeability of a randomized trial, Inverse Probability of Treatment Weighting (IPTW) was implemented using the Average Treatment Effect on the Overlap Population (ATO) weighting scheme ^17^. ATO weights were prioritized as they strictly bound the weights to a maximum of 1, thereby avoiding extreme variance inflation. The propensity scores were calculated via logistic regression incorporating age, sex, ethnicity, BMI, Townsend Deprivation Index, smoking status, alcohol frequency, standard cardiometabolic comorbidities, the AF PRS, and the top 10 genetic principal components. Covariate balance was confirmed by a maximum Absolute Standardized Mean Difference (ASMD) $< 0.1$

7.	Nielsen JB, Thorolfsdottir RB, Fritsche LG, Zhou W, Skov MW, Graham SE, Herron TJ, McCarthy S, Schmidt EM, Sveinbjornsson G, Surakka I, Mathis MR, Yamazaki M, Crawford RD, Gabrielsen ME, Skogholt AH, Holmen OL, Lin M, Wolford BN, Dey R, Dalen H, Sulem P, Chung JH, Backman JD, Arnar DO, Thorsteinsdottir U, Baras A, O'Dushlaine C, Holst AG, Wen X, Hornsby W, Dewey FE, Boehnke M, Kheterpal S, Mukherjee B, Lee S, Kang HM, Holm H, Kitzman J, Shavit JA, Jalife J, Brummett CM, Teslovich TM, Carey DJ, Gudbjartsson DF, Stefansson K, Abecasis GR, Hveem K, Willer CJ. Biobank-driven genomic discovery yields new insight into atrial fibrillation biology. *Nat Genet* 2018;**50**:1234–1239.

#### Supplementary Table S1. Baseline clinical characteristics of the study population stratified by Fibrosis-4 (FIB-4) index risk categories.

| **Characteristic** | **level** | **Overall** | **Low Risk** | **Intermediate Risk** | **High Risk** | **p** |
| --- | --- | --- | --- | --- | --- | --- |
| **n** |  | **403974** | **226289** | **168958** | **8727** |  |
| **Age at baseline, years (mean (SD))** |  | **56.48 (8.08)** | **53.37 (7.82)** | **60.34 (6.54)** | **62.22 (6.16)** | **<0.001** |
| **Sex (%)** | **Female** | **218560 (54.1)** | **133248 (58.9)** | **82258 (48.7)** | **3054 (35.0)** | **<0.001** |
|  | **Male** | **185414 (45.9)** | **93041 (41.1)** | **86700 (51.3)** | **5673 (65.0)** |  |
| **Race/Ethnicity (%)** | **White** | **369751 (91.5)** | **205766 (90.9)** | **156082 (92.4)** | **7903 (90.6)** | **<0.001** |
|  | **Asian** | **8614 (2.1)** | **5574 (2.5)** | **2876 (1.7)** | **164 (1.9)** |  |
|  | **Black** | **6138 (1.5)** | **3149 (1.4)** | **2741 (1.6)** | **248 (2.8)** |  |
|  | **Other/Mixed** | **19471 (4.8)** | **11800 (5.2)** | **7259 (4.3)** | **412 (4.7)** |  |
| **Body mass index, kg/m2 (mean (SD))** |  | **27.40 (4.76)** | **27.72 (4.92)** | **26.99 (4.50)** | **27.29 (4.85)** | **<0.001** |
| **Townsend Deprivation Index (mean (SD))** |  | **-1.32 (3.08)** | **-1.23 (3.12)** | **-1.46 (3.01)** | **-1.08 (3.24)** | **<0.001** |
| **Smoking status (%)** | **Current** | **42667 (10.6)** | **27533 (12.2)** | **14264 (8.5)** | **870 (10.0)** | **<0.001** |
|  | **Never** | **220211 (54.6)** | **125629 (55.6)** | **90287 (53.5)** | **4295 (49.3)** |  |
|  | **Prefer not to answer** | **1590 (0.4)** | **790 (0.3)** | **758 (0.4)** | **42 (0.5)** |  |
|  | **Previous** | **139118 (34.5)** | **72126 (31.9)** | **63484 (37.6)** | **3508 (40.3)** |  |
| **Diabetes mellitus (%)** |  | **23337 (5.8)** | **12562 (5.6)** | **9870 (5.8)** | **905 (10.4)** | **<0.001** |
| **Hypertension (%)** |  | **219267 (54.3)** | **111846 (49.4)** | **101443 (60.0)** | **5978 (68.5)** | **<0.001** |
| **Hyperlipidemia (%)** |  | **220913 (54.7)** | **124349 (55.0)** | **91525 (54.2)** | **5039 (57.7)** | **<0.001** |
| **Chronic kidney disease (%)** |  | **6157 (1.5)** | **2211 (1.0)** | **3625 (2.1)** | **321 (3.7)** | **<0.001** |
| **Cardiovascular disease (excl. AF) (%)** |  | **25316 (6.3)** | **10100 (4.5)** | **14086 (8.3)** | **1130 (12.9)** | **<0.001** |
| **NAFLD/MASLD (%)** |  | **154065 (38.2)** | **90342 (40.0)** | **60029 (35.6)** | **3694 (42.6)** | **<0.001** |
| **MetALD (%)** |  | **27520 (6.8)** | **15370 (6.8)** | **11386 (6.7)** | **764 (8.8)** | **<0.001** |
| **Viral hepatitis (%)** |  | **735 (0.2)** | **318 (0.1)** | **344 (0.2)** | **73 (0.8)** | **<0.001** |
| **ALT, U/L (median [IQR])** |  | **20.11 [15.38, 27.37]** | **20.39 [15.48, 27.87]** | **19.71 [15.27, 26.48]** | **21.62 [15.20, 34.08]** | **<0.001** |
| **AST, U/L (median [IQR])** |  | **24.30 [21.00, 28.80]** | **22.80 [19.80, 26.70]** | **26.20 [22.80, 30.80]** | **33.30 [27.00, 48.70]** | **<0.001** |
| **Triglycerides, mmol/L (median [IQR])** |  | **1.48 [1.04, 2.15]** | **1.52 [1.06, 2.21]** | **1.44 [1.03, 2.07]** | **1.38 [0.97, 2.03]** | **<0.001** |
| **Incident atrial fibrillation (%)** |  | **26677 (6.6)** | **10158 (4.5)** | **15246 (9.0)** | **1273 (14.6)** | **<0.001** |
| **Follow-up duration, years (median [IQR])** |  | **13.18 [12.31, 13.91]** | **13.31 [12.54, 14.02]** | **12.96 [12.14, 13.79]** | **12.53 [11.29, 13.49]** | **<0.001** |
| **PRS Tertile Group (%)** | **Low PRS** | **134658 (33.3)** | **75412 (33.3)** | **56383 (33.4)** | **2863 (32.8)** | 0.688 |
|  | **Intermediate PRS** | **134658 (33.3)** | **75498 (33.4)** | **56265 (33.3)** | **2895 (33.2)** |  |
|  | **High PRS** | **134658 (33.3)** | **75379 (33.3)** | **56310 (33.3)** | **2969 (34.0)** |  |

Continuous variables are presented as means (standard deviations, SD) for normally distributed data, or medians [interquartile ranges, IQR] for skewed data. Categorical variables are expressed as absolute frequencies (percentages). Group comparisons across the three risk strata (Low, Intermediate, and High Risk) were performed using Analysis of Variance (ANOVA) for normally distributed continuous variables, the Kruskal-Wallis test for non-normally distributed continuous variables, and Pearson’s chi-square test for categorical variables. Notably, while acquired cardiometabolic comorbidities were markedly enriched in the high-risk stratum, the innate genetic predisposition (PRS tertile groups) remained statistically balanced.

**Abbreviations:** AF, atrial fibrillation; ALT, alanine aminotransferase; AST, aspartate aminotransferase; BMI, body mass index; CKD, chronic kidney disease; CVD, cardiovascular disease; IQR, interquartile range; MASLD, metabolic dysfunction-associated steatotic liver disease; MetALD, metabolic dysfunction and alcohol-associated steatotic liver disease; NAFLD, non-alcoholic fatty liver disease; PRS, polygenic risk score; SD, standard deviation.

#### Supplementary Table S2. Baseline clinical characteristics of the study population stratified by NAFLD Fibrosis Score (NFS) risk categories.

| **Characteristic** | **level** | **Overall** | **Low Risk** | **Intermediate Risk** | **High Risk** | **p** |
| --- | --- | --- | --- | --- | --- | --- |
| **n** |  | 403974 | 284939 | 113930 | 5105 |  |
| **Age at baseline, years (mean (SD))** |  | 56.48 (8.08) | 54.74 (7.98) | 60.56 (6.73) | 62.00 (6.09) | <0.001 |
| **Sex (%)** | Female | 218560 (54.1) | 162697 (57.1) | 53683 (47.1) | 2180 (42.7) | <0.001 |
|  | Male | 185414 (45.9) | 122242 (42.9) | 60247 (52.9) | 2925 (57.3) |  |
| **Race/Ethnicity (%)** | White | 369751 (91.5) | 261458 (91.8) | 103851 (91.2) | 4442 (87.0) | <0.001 |
|  | Asian | 8614 (2.1) | 6214 (2.2) | 2256 (2.0) | 144 (2.8) |  |
|  | Black | 6138 (1.5) | 3195 (1.1) | 2713 (2.4) | 230 (4.5) |  |
|  | Other/Mixed | 19471 (4.8) | 14072 (4.9) | 5110 (4.5) | 289 (5.7) |  |
| **Body mass index, kg/m2 (mean (SD))** |  | 27.40 (4.76) | 26.47 (4.06) | 29.39 (5.26) | 35.02 (7.96) | <0.001 |
| **Townsend Deprivation Index (mean (SD))** |  | -1.32 (3.08) | -1.39 (3.05) | -1.20 (3.12) | -0.39 (3.44) | <0.001 |
| **Smoking status (%)** | Current | 42667 (10.6) | 31827 (11.2) | 10329 (9.1) | 511 (10.0) | <0.001 |
|  | Never | 220211 (54.6) | 159673 (56.1) | 58206 (51.2) | 2332 (45.7) |  |
|  | Prefer not to answer | 1590 (0.4) | 976 (0.3) | 575 (0.5) | 39 (0.8) |  |
|  | Previous | 139118 (34.5) | 92237 (32.4) | 44664 (39.3) | 2217 (43.5) |  |
| **Diabetes mellitus (%)** |  | 23337 (5.8) | 4529 (1.6) | 15831 (13.9) | 2977 (58.3) | <0.001 |
| **Hypertension (%)** |  | 219267 (54.3) | 140332 (49.2) | 74730 (65.6) | 4205 (82.4) | <0.001 |
| **Hyperlipidemia (%)** |  | 220913 (54.7) | 146067 (51.3) | 70811 (62.2) | 4035 (79.0) | <0.001 |
| **Chronic kidney disease (%)** |  | 6157 (1.5) | 2631 (0.9) | 3136 (2.8) | 390 (7.6) | <0.001 |
| **Cardiovascular disease (excl. AF) (%)** |  | 25316 (6.3) | 12349 (4.3) | 11865 (10.4) | 1102 (21.6) | <0.001 |
| **Incident atrial fibrillation (%)** |  | 26677 (6.6) | 13078 (4.6) | 12580 (11.0) | 1019 (20.0) | <0.001 |
| **Follow-up duration, years (median [IQR])** |  | 13.18 [12.31, 13.91] | 13.28 [12.51, 13.99] | 12.87 [12.06, 13.75] | 12.36 [9.22, 13.20] | <0.001 |
| **NAFLD/MASLD (%)** |  | 154065 (38.2) | 92261 (32.4) | 57914 (50.9) | 3890 (76.6) | <0.001 |
| **MetALD (%)** |  | 27520 (6.8) | 17673 (6.2) | 9372 (8.2) | 475 (9.3) | <0.001 |
| **Viral hepatitis (%)** |  | 735 (0.2) | 459 (0.2) | 246 (0.2) | 30 (0.6) | <0.001 |
| **ALT, U/L (median [IQR])** |  | 20.11 [15.38, 27.37] | 20.50 [15.72, 27.88] | 19.22 [14.67, 26.05] | 18.27 [11.69, 27.51] | <0.001 |
| **AST, U/L (median [IQR])** |  | 24.30 [21.00, 28.80] | 24.10 [20.80, 28.50] | 24.90 [21.50, 29.50] | 26.00 [21.60, 33.50] | <0.001 |
| **Triglycerides, mmol/L (median [IQR])** |  | 1.48 [1.04, 2.15] | 1.46 [1.03, 2.12] | 1.54 [1.09, 2.20] | 1.66 [1.17, 2.35] | <0.001 |
| **PRS Tertile Group (%)** | Low PRS | 134658 (33.3) | 95710 (33.6) | 37298 (32.7) | 1650 (32.3) | <0.001 |
|  | Intermediate PRS | 134658 (33.3) | 94972 (33.3) | 37965 (33.3) | 1721 (33.7) |  |
|  | High PRS | 134658 (33.3) | 94257 (33.1) | 38667 (33.9) | 1734 (34.0) |  |

Continuous variables are presented as means (standard deviations, SD) for normally distributed data, or medians [interquartile ranges, IQR] for skewed data. Categorical variables are expressed as absolute frequencies (percentages). Group comparisons across the three risk strata (Low, Intermediate, and High Risk) were performed using Analysis of Variance (ANOVA) for normally distributed continuous variables, the Kruskal-Wallis test for non-normally distributed continuous variables, and Pearson’s chi-square test for categorical variables. Notably, while acquired cardiometabolic comorbidities were markedly enriched in the high-risk stratum, the innate genetic predisposition (PRS tertile groups) remained statistically balanced.

**Abbreviations:** AF, atrial fibrillation; ALT, alanine aminotransferase; AST, aspartate aminotransferase; BMI, body mass index; CKD, chronic kidney disease; CVD, cardiovascular disease; IQR, interquartile range; MASLD, metabolic dysfunction-associated steatotic liver disease; MetALD, metabolic dysfunction and alcohol-associated steatotic liver disease; NAFLD, non-alcoholic fatty liver disease; PRS, polygenic risk score; SD, standard deviation.

#### Supplementary Table S3. Baseline clinical characteristics of the study population stratified by AST to Platelet Ratio Index (APRI) risk categories.

| **Characteristic** | **level** | **Overall** | **Low Risk** | **High Risk** | **p** |
| --- | --- | --- | --- | --- | --- |
| **n** |  | 403974 | 385788 | 18186 |  |
| **Age at baseline, years (mean (SD))** |  | 56.48 (8.08) | 56.44 (8.09) | 57.21 (7.96) | <0.001 |
| **Sex (%)** | Female | 218560 (54.1) | 213541 (55.4) | 5019 (27.6) | <0.001 |
|  | Male | 185414 (45.9) | 172247 (44.6) | 13167 (72.4) |  |
| **Race/Ethnicity (%)** | White | 369751 (91.5) | 353384 (91.6) | 16367 (90.0) | <0.001 |
|  | Asian | 8614 (2.1) | 8217 (2.1) | 397 (2.2) |  |
|  | Black | 6138 (1.5) | 5608 (1.5) | 530 (2.9) |  |
|  | Other/Mixed | 19471 (4.8) | 18579 (4.8) | 892 (4.9) |  |
| **Body mass index, kg/m2 (mean (SD))** |  | 27.40 (4.76) | 27.33 (4.73) | 28.94 (5.12) | <0.001 |
| **Townsend Deprivation Index (mean (SD))** |  | -1.32 (3.08) | -1.34 (3.07) | -0.93 (3.28) | <0.001 |
| **Smoking status (%)** | Current | 42667 (10.6) | 40678 (10.6) | 1989 (11.0) | <0.001 |
|  | Never | 220211 (54.6) | 211269 (54.8) | 8942 (49.2) |  |
|  | Prefer not to answer | 1590 (0.4) | 1502 (0.4) | 88 (0.5) |  |
|  | Previous | 139118 (34.5) | 131975 (34.2) | 7143 (39.3) |  |
| **Diabetes mellitus (%)** |  | 23337 (5.8) | 21010 (5.4) | 2327 (12.8) | <0.001 |
| **Hypertension (%)** |  | 219267 (54.3) | 206902 (53.6) | 12365 (68.0) | <0.001 |
| **Hyperlipidemia (%)** |  | 220913 (54.7) | 208710 (54.1) | 12203 (67.1) | <0.001 |
| **Chronic kidney disease (%)** |  | 6157 (1.5) | 5764 (1.5) | 393 (2.2) | <0.001 |
| **Cardiovascular disease (excl. AF) (%)** |  | 25316 (6.3) | 23326 (6.0) | 1990 (10.9) | <0.001 |
| **Incident atrial fibrillation (%)** |  | 26677 (6.6) | 24819 (6.4) | 1858 (10.2) | <0.001 |
| **Follow-up duration, years (median [IQR])** |  | 13.18 [12.31, 13.91] | 13.19 [12.32, 13.92] | 12.83 [12.03, 13.74] | <0.001 |
| **NAFLD/MASLD (%)** |  | 154065 (38.2) | 142521 (37.0) | 11544 (63.8) | <0.001 |
| **MetALD (%)** |  | 27520 (6.8) | 24946 (6.5) | 2574 (14.2) | <0.001 |
| **Viral hepatitis (%)** |  | 735 (0.2) | 581 (0.2) | 154 (0.8) | <0.001 |
| **ALT, U/L (median [IQR])** |  | 20.11 [15.38, 27.37] | 19.71 [15.20, 26.37] | 43.14 [28.91, 63.95] | <0.001 |
| **AST, U/L (median [IQR])** |  | 24.30 [21.00, 28.80] | 24.10 [20.80, 28.10] | 44.50 [35.60, 57.30] | <0.001 |
| **Triglycerides, mmol/L (median [IQR])** |  | 1.48 [1.04, 2.15] | 1.47 [1.04, 2.13] | 1.70 [1.13, 2.56] | <0.001 |
| **PRS Tertile Group (%)** | Low PRS | 134658 (33.3) | 128630 (33.3) | 6028 (33.1) | 0.840 |
|  | Intermediate PRS | 134658 (33.3) | 128591 (33.3) | 6067 (33.4) |  |
|  | High PRS | 134658 (33.3) | 128567 (33.3) | 6091 (33.5) |  |

Continuous variables are presented as means (standard deviations, SD) for normally distributed data, or medians [interquartile ranges, IQR] for skewed data. Categorical variables are expressed as absolute frequencies (percentages). Group comparisons across the three risk strata (Low, Intermediate, and High Risk) were performed using Analysis of Variance (ANOVA) for normally distributed continuous variables, the Kruskal-Wallis test for non-normally distributed continuous variables, and Pearson’s chi-square test for categorical variables. Notably, while acquired cardiometabolic comorbidities were markedly enriched in the high-risk stratum, the innate genetic predisposition (PRS tertile groups) remained statistically balanced.

**Abbreviations:** AF, atrial fibrillation; ALT, alanine aminotransferase; AST, aspartate aminotransferase; BMI, body mass index; CKD, chronic kidney disease; CVD, cardiovascular disease; IQR, interquartile range; MASLD, metabolic dysfunction-associated steatotic liver disease; MetALD, metabolic dysfunction and alcohol-associated steatotic liver disease; NAFLD, non-alcoholic fatty liver disease; PRS, polygenic risk score; SD, standard deviation.

#### Supplementary Table S4. Baseline clinical characteristics of the study population stratified by AST/ALT ratio risk categories.

| **Characteristic** | **level** | **Overall** | **Low (<1)** | **Intermediate**  **(1-2)** | **High (>2)** | **p** |
| --- | --- | --- | --- | --- | --- | --- |
| **n** |  | 403974 | 112114 | 273562 | 18298 |  |
| **Age at baseline, years (mean (SD))** |  | 56.48 (8.08) | 55.52 (7.88) | 56.86 (8.11) | 56.59 (8.44) | <0.001 |
| **Sex (%)** | Female | 218560 (54.1) | 39631 (35.3) | 166172 (60.7) | 12757 (69.7) | <0.001 |
|  | Male | 185414 (45.9) | 72483 (64.7) | 107390 (39.3) | 5541 (30.3) |  |
| **Race/Ethnicity (%)** | White | 369751 (91.5) | 102840 (91.7) | 250626 (91.6) | 16285 (89.0) | <0.001 |
|  | Asian | 8614 (2.1) | 2676 (2.4) | 5488 (2.0) | 450 (2.5) |  |
|  | Black | 6138 (1.5) | 1242 (1.1) | 4395 (1.6) | 501 (2.7) |  |
|  | Other/Mixed | 19471 (4.8) | 5356 (4.8) | 13053 (4.8) | 1062 (5.8) |  |
| **Body mass index, kg/m2 (mean (SD))** |  | 27.40 (4.76) | 29.51 (4.76) | 26.68 (4.51) | 25.32 (4.30) | <0.001 |
| **Townsend Deprivation Index (mean (SD))** |  | -1.32 (3.08) | -1.26 (3.11) | -1.36 (3.06) | -1.18 (3.15) | <0.001 |
| **Smoking status (%)** | Current | 42667 (10.6) | 12526 (11.2) | 28112 (10.3) | 2029 (11.1) | <0.001 |
|  | Never | 220211 (54.6) | 57794 (51.6) | 152088 (55.6) | 10329 (56.6) |  |
|  | Prefer not to answer | 1590 (0.4) | 466 (0.4) | 1050 (0.4) | 74 (0.4) |  |
|  | Previous | 139118 (34.5) | 41225 (36.8) | 92060 (33.7) | 5833 (31.9) |  |
| **Diabetes mellitus (%)** |  | 23337 (5.8) | 11637 (10.4) | 11167 (4.1) | 533 (2.9) | <0.001 |
| **Hypertension (%)** |  | 219267 (54.3) | 70929 (63.3) | 139945 (51.2) | 8393 (45.9) | <0.001 |
| **Hyperlipidemia (%)** |  | 220913 (54.7) | 81713 (72.9) | 132543 (48.5) | 6657 (36.4) | <0.001 |
| **Chronic kidney disease (%)** |  | 6157 (1.5) | 1318 (1.2) | 4414 (1.6) | 425 (2.3) | <0.001 |
| **Cardiovascular disease (excl. AF) (%)** |  | 25316 (6.3) | 8089 (7.2) | 16228 (5.9) | 999 (5.5) | <0.001 |
| **NAFLD/MASLD (%)** |  | 154065 (38.2) | 75607 (67.5) | 75639 (27.7) | 2819 (15.4) | <0.001 |
| **MetALD (%)** |  | 27520 (6.8) | 14314 (12.8) | 12757 (4.7) | 449 (2.5) | <0.001 |
| **Viral hepatitis (%)** |  | 735 (0.2) | 236 (0.2) | 444 (0.2) | 55 (0.3) | <0.001 |
| **ALT, U/L (median [IQR])** |  | 20.11 [15.38, 27.37] | 31.91 [25.96, 41.08] | 17.86 [14.63, 21.91] | 10.25 [8.47, 12.47] | <0.001 |
| **AST, U/L (median [IQR])** |  | 24.30 [21.00, 28.80] | 26.60 [22.50, 32.10] | 23.70 [20.50, 27.50] | 23.90 [20.60, 28.40] | <0.001 |
| **Triglycerides, mmol/L (median [IQR])** |  | 1.48 [1.04, 2.15] | 1.90 [1.34, 2.69] | 1.36 [0.98, 1.94] | 1.16 [0.86, 1.62] | <0.001 |
| **Incident atrial fibrillation (%)** |  | 26677 (6.6) | 7309 (6.5) | 18173 (6.6) | 1195 (6.5) | 0.343 |
| **Follow-up duration, years (median [IQR])** |  | 13.18 [12.31, 13.91] | 13.22 [12.32, 14.02] | 13.18 [12.31, 13.89] | 12.85 [12.24, 13.66] | <0.001 |
| **PRS Tertile Group (%)** | Low PRS | 134658 (33.3) | 37151 (33.1) | 91250 (33.4) | 6257 (34.2) | 0.061 |
|  | Intermediate PRS | 134658 (33.3) | 37471 (33.4) | 91116 (33.3) | 6071 (33.2) |  |
|  | High PRS | 134658 (33.3) | 37492 (33.4) | 91196 (33.3) | 5970 (32.6) |  |

Continuous variables are presented as means (standard deviations, SD) for normally distributed data, or medians [interquartile ranges, IQR] for skewed data. Categorical variables are expressed as absolute frequencies (percentages). Group comparisons across the three risk strata (Low, Intermediate, and High Risk) were performed using Analysis of Variance (ANOVA) for normally distributed continuous variables, the Kruskal-Wallis test for non-normally distributed continuous variables, and Pearson’s chi-square test for categorical variables. Notably, while acquired cardiometabolic comorbidities were markedly enriched in the high-risk stratum, the innate genetic predisposition (PRS tertile groups) remained statistically balanced.

**Abbreviations:** AF, atrial fibrillation; ALT, alanine aminotransferase; AST, aspartate aminotransferase; BMI, body mass index; CKD, chronic kidney disease; CVD, cardiovascular disease; IQR, interquartile range; MASLD, metabolic dysfunction-associated steatotic liver disease; MetALD, metabolic dysfunction and alcohol-associated steatotic liver disease; NAFLD, non-alcoholic fatty liver disease; PRS, polygenic risk score; SD, standard deviation.

#### Supplementary Table S5. Baseline clinical characteristics of the imaging subcohort stratified by liver corrected T1 (cT1) risk categories.

| **Characteristic** | **level** | **Overall** | **Low Risk** | **High Risk** | **p** |
| --- | --- | --- | --- | --- | --- |
| **n** |  | 27032 | 22638 | 4394 |  |
| **Age at MRI scan, years (median [IQR])** |  | 64.61 [58.22, 70.16] | 64.55 [58.23, 70.14] | 64.87 [58.17, 70.25] | 0.357 |
| **Sex (%)** | Female | 14216 (52.6) | 12379 (54.7) | 1837 (41.8) | **<0.001** |
|  | Male | 12816 (47.4) | 10259 (45.3) | 2557 (58.2) |  |
| **Race/Ethnicity (%)** | White | 25348 (93.8) | 21223 (93.7) | 4125 (93.9) | 0.340 |
|  | Asian | 360 (1.3) | 294 (1.3) | 66 (1.5) |  |
|  | Black | 174 (0.6) | 142 (0.6) | 32 (0.7) |  |
|  | Other/Mixed | 1150 (4.3) | 979 (4.3) | 171 (3.9) |  |
| **Body mass index (MRI), kg/m2 (median [IQR])** |  | 25.79 [23.41, 28.69] | 25.30 [23.09, 27.80] | 29.34 [26.29, 32.59] | **<0.001** |
| **Townsend Deprivation Index (median [IQR])** |  | -2.61 [-3.88, -0.46] | -2.64 [-3.90, -0.53] | -2.41 [-3.77, 0.00] | **<0.001** |
| **Smoking status (%)** | Current | 865 (3.2) | 684 (3.0) | 181 (4.1) | **<0.001** |
|  | Never | 16905 (62.9) | 14340 (63.7) | 2565 (58.8) |  |
|  | Prefer not to answer | 85 (0.3) | 73 (0.3) | 12 (0.3) |  |
|  | Previous | 9025 (33.6) | 7421 (33.0) | 1604 (36.8) |  |
| **Diabetes mellitus (%)** |  | 1397 (5.2) | 805 (3.6) | 592 (13.5) | **<0.001** |
| **Hypertension (%)** |  | 13207 (48.9) | 10445 (46.1) | 2762 (62.9) | **<0.001** |
| **Hyperlipidemia (%)** |  | 14323 (53.0) | 11006 (48.6) | 3317 (75.5) | **<0.001** |
| **Chronic kidney disease (%)** |  | 308 (1.1) | 239 (1.1) | 69 (1.6) | **0.004** |
| **CVD History (excl. AF) (%)** |  | 1879 (7.0) | 1421 (6.3) | 458 (10.4) | **<0.001** |
| **NAFLD/MASLD (%)** |  | 7343 (27.2) | 4281 (18.9) | 3062 (69.7) | **<0.001** |
| **MetALD (%)** |  | 2014 (7.5) | 1341 (5.9) | 673 (15.3) | **<0.001** |
| **Viral hepatitis (%)** |  | 134 (0.5) | 113 (0.5) | 21 (0.5) | 0.947 |
| **Liver PDFF, % (median [IQR])** |  | 3.10 [2.20, 5.40] | 2.80 [2.10, 4.30] | 9.40 [4.20, 15.90] | **<0.001** |
| **Incident AF (Post-MRI) (%)** |  | 500 (1.8) | 374 (1.7) | 126 (2.9) | **<0.001** |
| **Follow-up duration, years (median [IQR])** |  | 3.71 [3.04, 4.67] | 3.71 [3.03, 4.67] | 3.74 [3.05, 4.70] | 0.145 |
| **PRS Tertile Group (%)** | Low PRS | 9011 (33.3) | 7570 (33.4) | 1441 (32.8) | 0.097 |
|  | Intermediate PRS | 9011 (33.3) | 7485 (33.1) | 1526 (34.7) |  |
|  | High PRS | 9010 (33.3) | 7583 (33.5) | 1427 (32.5) |  |

Continuous variables are presented as medians [interquartile ranges, IQR], and categorical variables are expressed as absolute frequencies (percentages). Group comparisons between the low-risk (cT1 < 750 ms) and high-risk (cT1 $\geq$ 750 ms) fibro-inflammation strata were performed using the Mann-Whitney U test for continuous variables and Pearson’s chi-square test for categorical variables. Consistent with the primary cohort findings, the high-risk cT1 group exhibited a markedly heavier burden of acquired cardiometabolic derangements—including severe concurrent hepatic steatosis (significantly elevated Liver PDFF)—yet maintained a comparable innate genetic predisposition (PRS tertile groups, $\text{P}\text{=0.097}$) and demographic baseline (e.g., age and race) relative to the low-risk group. **Abbreviations:** AF, atrial fibrillation; BMI, body mass index; CKD, chronic kidney disease; cT1, corrected T1; CVD, cardiovascular disease; IQR, interquartile range; MASLD, metabolic dysfunction-associated steatotic liver disease; Met-ALD, metabolic dysfunction and alcohol-associated steatotic liver disease; MRI, magnetic resonance imaging; NAFLD, non-alcoholic fatty liver disease; PDFF, proton density fat fraction; PRS, polygenic risk score; TDI, Townsend Deprivation Index.

#### Supplementary Table S6. Comprehensive sensitivity analyses for the association between liver fibrosis indices and incident atrial fibrillation.

| **HR (95% CI)**  **P value**  **(Ref : Low Risk)**  **Sensitivity Scenario** | **FIB-4 High Risk** | **NFS High Risk** | **APRI High Risk** | **AST/ALT High Risk** | **cT - 1 High Risk** |
| --- | --- | --- | --- | --- | --- |
| Excluding incident AF cases diagnosed within the first two years of follow-up | 1.48 (1.39-1.58)  P <0.001 | 1.48 (1.37-1.60)  P <0.001 | 1.18 (1.13-1.24)  P <0.001 | 1.43 (1.34-1.53)  P <0.001 | - |
| Excluding participants with pre-existing cardiovascular disease at baseline. | 1.55 (1.45-1.66)  P <0.001 | 1.57 (1.44-1.72)  P <0.001 | 1.21 (1.15-1.28)  P <0.001 | 1.43 (1.34-1.54)  P <0.001 | - |
| Excluding former or harmful drinkers. | 1.47 (1.38-1.57)  P <0.001 | 1.51 (1.40-1.64)  P <0.001 | 1.17 (1.11-1.23)  P <0.001 | 1.41 (1.32-1.51)  P <0.001 | - |
| Excluding participants with extreme liver enzyme elevations or hematological malignancies. | 1.51 (1.42-1.61)  P <0.001 | 1.53 (1.42-1.64)  P <0.001 | 1.20 (1.14-1.26)  P <0.001 | 1.44 (1.35-1.53)  P <0.001 | - |
| Utilizing a stringent definition of incident atrial fibrillation (strictly excluding atrial flutter). | 1.54 (1.44-1.64)  P <0.001 | 1.50 (1.38-1.63)  P <0.001 | 1.12 (1.10-1.14)  P <0.001 | 1.47 (1.37-1.57)  P <0.001 | - |
| Applying alternative clinical thresholds and reclassification schemes for biochemical liver fibrosis indices. | 1.46 (1.38-1.55)  P <0.001 | 1.41 (1.31-1.51)  P <0.001 | 1.44 (1.27-1.64)  P <0.001 | 1.29 (1.25-1.33)  P <0.001 | - |
| Using Fine-Gray subdistribution hazard models to account for the competing risk of mortality | 1.44 (1.36-1.53)  P <0.001 | 1.44 (1.34-1.55)  P <0.001 | 1.17 (1.11-1.22)  P <0.001 | 1.37 (1.29-1.46)  P <0.001 | 1.37 (1.07-1.76)  P =0.013 |
| Using Inverse Probability of Treatment Weighting (IPTW) in the primary cohort. | 1.53 (1.43-1.63)  P <0.001 | 1.77 (1.62-1.93)  P <0.001 | 1.19 (1.14-1.25)  P <0.001 | 1.35 (1.26-1.45)  P <0.001 | 1.36 (1.06-1.74)  P =0.016 |

Hazard ratios (HRs) and 95% confidence intervals (CIs) for incident atrial fibrillation were estimated for the high-risk categories of each liver fibrosis index compared to their respective low-risk reference groups (Ref). Unless otherwise specified, all scenarios utilized multivariable Cox proportional hazards models comprehensively adjusted for age, sex, ethnicity, body mass index, smoking status, alcohol intake frequency, Townsend deprivation index, hypertension, type 2 diabetes mellitus, lipid-lowering medication use, chronic kidney disease, prevalent cardiovascular disease, the standardized polygenic risk score for atrial fibrillation, and the first 10 genetic principal components (equivalent to the Fully Adjusted Model 4). For the competing risk scenario, Fine-Gray models were applied, and estimates represent subdistribution hazard ratios (sHRs). For the Inverse Probability of Treatment Weighting (IPTW) scenario, Average Treatment Effect on the Overlap Population (ATO) weights were used to balance all aforementioned baseline covariates. Missing estimates (-) for cT-1 indicate scenarios that were not applicable or not performed within the imaging subcohort. **Abbreviations:** AF, atrial fibrillation; APRI, AST to Platelet Ratio Index; AST/ALT, aspartate aminotransferase to alanine aminotransferase ratio; CI, confidence interval; cT1, liver corrected T1; FIB-4, Fibrosis-4 index; HR, hazard ratio; IPTW, Inverse Probability of Treatment Weighting; NFS, NAFLD fibrosis score; Ref, reference.

#### Supplementary Table S7. Detailed causal mediation estimates across the core liver-heart metabolome.

| **Liver Fibrosis Index** | **Metabolite Class** | **Metabolite** | **Total Effect**  **(HR, 95% CI)** | **Total Effect P-value** | **ACME**  **(HR, 95% CI)** | **ACME P-value** | **Proportion**  **(%)** |
| --- | --- | --- | --- | --- | --- | --- | --- |
| FIB-4 | Apolipoproteins | Apolipoprotein.B | 1.118 (1.104-1.132) | <0.001 | 1.010 (1.009-1.012) | <0.001 | 9.5% (7.8%, 11.2%) |
|  |  | Ratio.of.apolipoprotein.B.to.apolipoprotein.A1 | 1.117 (1.103-1.131) | <0.001 | 1.007 (1.006-1.009) | <0.001 | 7.0% (5.6%, 8.5%) |
|  | Cholesterol | VLDL.cholesterol | 1.118 (1.104-1.133) | <0.001 | 1.011 (1.010-1.013) | <0.001 | 10.6% (8.8%, 12.3%) |
|  |  | LDL.cholesterol | 1.117 (1.103-1.132) | <0.001 | 1.011 (1.009-1.012) | <0.001 | 10.1% (8.4%, 11.9%) |
|  |  | Total.cholesterol ★ | 1.117 (1.103-1.131) | <0.001 | 1.006 (1.005-1.008) | <0.001 | 6.1% (4.8%, 7.4%) |
|  | Cholesteryl esters | Cholesteryl.esters.in.LDL | 1.118 (1.104-1.132) | <0.001 | 1.012 (1.010-1.013) | <0.001 | 11.1% (9.2%, 12.9%) |
|  |  | Cholesteryl.esters.in.VLDL | 1.118 (1.104-1.132) | <0.001 | 1.010 (1.008-1.011) | <0.001 | 9.2% (7.6%, 10.9%) |
|  | Fatty acids | Total.fatty.acids | 1.117 (1.103-1.132) | <0.001 | 1.009 (1.007-1.010) | <0.001 | 8.1% (6.7%, 9.6%) |
|  |  | Monounsaturated.fatty.acids | 1.117 (1.103-1.132) | <0.001 | 1.008 (1.007-1.010) | <0.001 | 7.8% (6.4%, 9.2%) |
|  |  | Linoleic.acid ★ | 1.117 (1.103-1.131) | <0.001 | 1.008 (1.006-1.009) | <0.001 | 7.2% (5.9%, 8.5%) |
|  |  | Ratio.of.monounsaturated.fatty.acids.to.total.fatty.acids ★ | 1.116 (1.102-1.131) | <0.001 | 1.004 (1.003-1.005) | <0.001 | 3.4% (2.4%, 4.4%) |
|  |  | Ratio.of.polyunsaturated.fatty.acids.to.monounsaturated.fatty.acids ★ | 1.116 (1.102-1.131) | <0.001 | 1.003 (1.002-1.004) | <0.001 | 3.1% (2.1%, 4.1%) |
|  | Free cholesterol | Free.cholesterol.in.VLDL | 1.119 (1.104-1.133) | <0.001 | 1.012 (1.011-1.014) | <0.001 | 11.6% (9.8%, 13.5%) |
|  |  | Free.cholesterol.in.LDL | 1.117 (1.103-1.131) | <0.001 | 1.008 (1.007-1.009) | <0.001 | 7.7% (6.1%, 9.2%) |
|  | Inflammation | Glycoprotein.acetyls ★ | 1.116 (1.102-1.131) | <0.001 | 1.007 (1.004-1.010) | <0.001 | 6.6% (3.6%, 9.6%) |
|  | Ketone bodies | Acetone ★ | 1.116 (1.102-1.130) | <0.001 | 1.004 (1.003-1.005) | <0.001 | 4.3% (3.2%, 5.4%) |
|  | Lipoprotein particle concentrations | Concentration.of.VLDL.particles | 1.118 (1.104-1.133) | <0.001 | 1.011 (1.009-1.012) | <0.001 | 10.0% (8.3%, 11.7%) |
|  |  | Concentration.of.LDL.particles | 1.118 (1.104-1.132) | <0.001 | 1.011 (1.009-1.012) | <0.001 | 9.9% (8.2%, 11.6%) |
|  | Lipoprotein particle sizes | Average.diameter.for.HDL.particles | 1.118 (1.104-1.132) | <0.001 | 1.013 (1.011-1.015) | <0.001 | 12.2% (10.1%, 14.3%) |
|  |  | Average.diameter.for.VLDL.particles | 1.117 (1.103-1.132) | <0.001 | 1.013 (1.011-1.014) | <0.001 | 12.0% (10.2%, 13.8%) |
|  | Lipoprotein subclasses | Total.lipids.in.medium.VLDL | 1.119 (1.105-1.133) | <0.001 | 1.015 (1.014-1.017) | <0.001 | 14.2% (12.1%, 16.4%) |
|  |  | Cholesteryl.esters.in.medium.LDL | 1.119 (1.104-1.133) | <0.001 | 1.015 (1.013-1.017) | <0.001 | 13.9% (11.8%, 16.1%) |
|  |  | Cholesteryl.esters.in.medium.VLDL ★ | 1.117 (1.103-1.131) | <0.001 | 1.006 (1.005-1.008) | <0.001 | 6.0% (4.7%, 7.3%) |
|  | Other lipids | Ratio.of.triglycerides.to.phosphoglycerides | 1.117 (1.103-1.132) | <0.001 | 1.009 (1.007-1.010) | <0.001 | 8.1% (6.7%, 9.5%) |
|  | Phospholipids | Phospholipids.in.VLDL | 1.119 (1.104-1.133) | <0.001 | 1.012 (1.011-1.014) | <0.001 | 11.4% (9.6%, 13.2%) |
|  |  | Phospholipids.in.LDL | 1.118 (1.104-1.132) | <0.001 | 1.011 (1.010-1.013) | <0.001 | 10.6% (8.7%, 12.4%) |
|  | Relative lipoprotein lipid concentrations | Triglycerides.to.total.lipids.ratio.in.very.large.HDL | 1.117 (1.103-1.132) | <0.001 | 1.008 (1.007-1.010) | <0.001 | 8.0% (6.6%, 9.3%) |
|  |  | Phospholipids.to.total.lipids.ratio.in.small.LDL | 1.117 (1.103-1.131) | <0.001 | 1.007 (1.006-1.008) | <0.001 | 6.4% (5.2%, 7.5%) |
|  |  | Triglycerides.to.total.lipids.ratio.in.small.HDL ★ | 1.117 (1.103-1.131) | <0.001 | 1.004 (1.003-1.005) | <0.001 | 4.0% (3.1%, 4.9%) |
|  | Total lipids | Total.lipids.in.VLDL | 1.119 (1.104-1.133) | <0.001 | 1.014 (1.012-1.015) | <0.001 | 12.7% (10.8%, 14.6%) |
|  |  | Total.lipids.in.LDL | 1.117 (1.103-1.132) | <0.001 | 1.011 (1.009-1.013) | <0.001 | 10.3% (8.5%, 12.1%) |
|  | Triglycerides | Triglycerides.in.VLDL | 1.118 (1.104-1.132) | <0.001 | 1.012 (1.011-1.013) | <0.001 | 11.3% (9.5%, 13.0%) |
|  |  | Total.triglycerides | 1.118 (1.104-1.132) | <0.001 | 1.011 (1.010-1.012) | <0.001 | 10.3% (8.6%, 11.9%) |
| NFS | Apolipoproteins | Apolipoprotein.B | 1.167 (1.148-1.186) | <0.001 | 1.014 (1.012-1.016) | <0.001 | 9.7% (8.0%, 11.3%) |
|  |  | Ratio.of.apolipoprotein.B.to.apolipoprotein.A1 | 1.166 (1.148-1.186) | <0.001 | 1.008 (1.006-1.009) | <0.001 | 5.4% (4.3%, 6.5%) |
|  | Cholesterol | VLDL.cholesterol | 1.167 (1.148-1.186) | <0.001 | 1.016 (1.014-1.018) | <0.001 | 11.0% (9.2%, 12.8%) |
|  |  | LDL.cholesterol | 1.167 (1.148-1.186) | <0.001 | 1.015 (1.013-1.017) | <0.001 | 10.3% (8.5%, 12.1%) |
|  |  | Total.cholesterol ★ | 1.166 (1.147-1.185) | <0.001 | 1.010 (1.008-1.012) | <0.001 | 7.1% (5.6%, 8.6%) |
|  | Cholesteryl esters | Cholesteryl.esters.in.LDL | 1.167 (1.148-1.186) | <0.001 | 1.017 (1.014-1.019) | <0.001 | 11.4% (9.5%, 13.3%) |
|  |  | Cholesteryl.esters.in.VLDL | 1.167 (1.148-1.187) | <0.001 | 1.014 (1.012-1.016) | <0.001 | 9.6% (7.9%, 11.3%) |
|  | Fatty acids | Total.fatty.acids | 1.166 (1.147-1.185) | <0.001 | 1.015 (1.013-1.017) | <0.001 | 10.3% (8.5%, 12.2%) |
|  |  | Polyunsaturated.fatty.acids | 1.167 (1.148-1.186) | <0.001 | 1.014 (1.012-1.016) | <0.001 | 9.6% (8.0%, 11.3%) |
|  |  | Linoleic.acid ★ | 1.167 (1.148-1.186) | <0.001 | 1.011 (1.010-1.013) | <0.001 | 7.9% (6.5%, 9.3%) |
|  |  | Ratio.of.monounsaturated.fatty.acids.to.total.fatty.acids ★ | 1.166 (1.147-1.185) | <0.001 | 1.005 (1.003-1.006) | <0.001 | 3.4% (2.3%, 4.4%) |
|  |  | Ratio.of.polyunsaturated.fatty.acids.to.monounsaturated.fatty.acids ★ | 1.165 (1.147-1.185) | <0.001 | 1.005 (1.003-1.006) | <0.001 | 3.2% (2.0%, 4.3%) |
|  | Free cholesterol | Free.cholesterol.in.VLDL | 1.167 (1.148-1.186) | <0.001 | 1.018 (1.016-1.020) | <0.001 | 12.2% (10.3%, 14.1%) |
|  |  | Total.free.cholesterol | 1.167 (1.148-1.186) | <0.001 | 1.012 (1.010-1.014) | <0.001 | 8.1% (6.5%, 9.7%) |
|  | Inflammation | Glycoprotein.acetyls ★ | 1.166 (1.147-1.185) | <0.001 | 1.007 (1.003-1.011) | <0.001 | 5.0% (2.2%, 7.8%) |
|  | Ketone bodies | Acetone ★ | 1.165 (1.146-1.184) | <0.001 | 1.003 (1.002-1.004) | <0.001 | 2.1% (1.6%, 2.6%) |
|  | Lipoprotein particle concentrations | Concentration.of.VLDL.particles | 1.167 (1.148-1.186) | <0.001 | 1.016 (1.014-1.018) | <0.001 | 11.0% (9.2%, 12.8%) |
|  |  | Concentration.of.LDL.particles | 1.167 (1.148-1.187) | <0.001 | 1.015 (1.012-1.017) | <0.001 | 10.1% (8.3%, 11.8%) |
|  | Lipoprotein particle sizes | Average.diameter.for.VLDL.particles | 1.166 (1.147-1.185) | <0.001 | 1.014 (1.013-1.016) | <0.001 | 10.0% (8.6%, 11.5%) |
|  |  | Average.diameter.for.HDL.particles | 1.166 (1.147-1.185) | <0.001 | 1.013 (1.011-1.015) | <0.001 | 8.9% (7.4%, 10.3%) |
|  | Lipoprotein subclasses | Cholesteryl.esters.in.medium.LDL | 1.167 (1.148-1.187) | <0.001 | 1.021 (1.018-1.023) | <0.001 | 14.3% (12.1%, 16.5%) |
|  |  | Total.lipids.in.medium.VLDL | 1.167 (1.148-1.187) | <0.001 | 1.020 (1.018-1.023) | <0.001 | 14.0% (11.9%, 16.0%) |
|  |  | Cholesteryl.esters.in.medium.VLDL ★ | 1.167 (1.148-1.186) | <0.001 | 1.009 (1.007-1.010) | <0.001 | 5.9% (4.6%, 7.2%) |
|  | Other lipids | Ratio.of.triglycerides.to.phosphoglycerides | 1.166 (1.147-1.185) | <0.001 | 1.010 (1.008-1.011) | <0.001 | 6.8% (5.6%, 7.9%) |
|  | Phospholipids | Phospholipids.in.VLDL | 1.167 (1.148-1.186) | <0.001 | 1.018 (1.015-1.020) | <0.001 | 12.1% (10.2%, 13.9%) |
|  |  | Phospholipids.in.LDL | 1.167 (1.148-1.186) | <0.001 | 1.016 (1.013-1.018) | <0.001 | 10.7% (8.9%, 12.6%) |
|  | Relative lipoprotein lipid concentrations | Triglycerides.to.total.lipids.ratio.in.very.large.HDL | 1.165 (1.146-1.184) | <0.001 | 1.013 (1.011-1.015) | <0.001 | 8.9% (7.4%, 10.4%) |
|  |  | Phospholipids.to.total.lipids.ratio.in.small.LDL | 1.166 (1.147-1.185) | <0.001 | 1.011 (1.010-1.013) | <0.001 | 8.0% (6.5%, 9.4%) |
|  |  | Triglycerides.to.total.lipids.ratio.in.small.HDL ★ | 1.166 (1.147-1.185) | <0.001 | 1.005 (1.004-1.006) | <0.001 | 3.7% (2.8%, 4.5%) |
|  | Total lipids | Total.lipids.in.VLDL | 1.167 (1.148-1.186) | <0.001 | 1.019 (1.017-1.021) | <0.001 | 12.9% (11.0%, 14.8%) |
|  |  | Total.lipids.in.LDL | 1.167 (1.148-1.186) | <0.001 | 1.015 (1.013-1.018) | <0.001 | 10.7% (8.8%, 12.5%) |
|  | Triglycerides | Triglycerides.in.VLDL | 1.166 (1.147-1.185) | <0.001 | 1.016 (1.014-1.018) | <0.001 | 11.2% (9.5%, 12.9%) |
|  |  | Total.triglycerides | 1.166 (1.147-1.185) | <0.001 | 1.016 (1.014-1.018) | <0.001 | 10.9% (9.2%, 12.6%) |

Total Effect (TE) and Average Causal Mediation Effect (ACME) are expressed as HRs per 1-standard deviation (SD) increment of the metabolite. The table features top mediating metabolites (mediated proportion >= 5%) alongside ultra-stable mediators. The star symbol (**★**) denotes ultra-stable core metabolites identified via Elastic Net penalization. Models were adjusted for baseline clinical covariates and genetic factors. **Abbreviations:** ACME, Average Causal Mediation Effect; CI, confidence interval; FIB-4, Fibrosis-4 index; HR, hazard ratio; NFS, NAFLD fibrosis score; TE, Total Effect.

#### Supplementary Table S8. Baseline clinical characteristics stratified by machine-learning-derived systemic metabolic phenotypes.

| **Characteristics** | **level** | **Healthy** | **Fibro-Inflammatory** | **High-Risk Comorbid** | **P value** |
| --- | --- | --- | --- | --- | --- |
| **n** |  | **217256** | **29659** | **115135** |  |
| Age (years), mean (SD) |  | 56.00 (8.05) | 56.98 (7.93) | 57.36 (8.11) | **<0.001** |
| Male (%) |  | 87934 (40.5) | 11565 (39.0) | 70031 (60.8) | **<0.001** |
| BMI (kg/m2), mean (SD) |  | 26.85 (4.44) | 26.10 (4.54) | 29.07 (4.99) | **<0.001** |
| Townsend Deprivation Index, mean (SD) |  | -1.48 (2.99) | -1.27 (3.08) | -1.08 (3.21) | **<0.001** |
| AST (U/L), median (IQR) |  | 23.90 [20.70, 27.90] | 24.50 [21.10, 29.10] | 25.40 [21.70, 30.40] | **<0.001** |
| ALT (U/L), median (IQR) |  | 19.04 [14.82, 25.36] | 18.21 [14.27, 24.27] | 23.52 [17.71, 32.20] | **<0.001** |
| AST/ALT Ratio, median (IQR) |  | 1.24 [1.01, 1.50] | 1.34 [1.10, 1.61] | 1.07 [0.88, 1.32] | **<0.001** |
| Platelet Count (10^9/L), mean (SD) |  | 254.65 (57.74) | 250.68 (58.95) | 252.56 (63.83) | **<0.001** |
| Hypertension, n (%) |  | 106902 (49.2) | 15850 (53.4) | 76145 (66.1) | **<0.001** |
| Type 2 Diabetes, n (%) |  | 4612 (2.1) | 1041 (3.5) | 14854 (12.9) | **<0.001** |
| Dyslipidemia, n (%) |  | 91109 (41.9) | 9388 (31.7) | 101260 (87.9) | **<0.001** |
| Chronic Kidney Disease, n (%) |  | 2160 (1.0) | 404 (1.4) | 2973 (2.6) | **<0.001** |
| Cardiovascular Disease, n (%) |  | 7003 (3.2) | 1545 (5.2) | 13872 (12.0) | **<0.001** |
| Incident AF (Follow-up), n (%) |  | 11859 (5.5) | 2110 (7.1) | 10026 (8.7) | **<0.001** |
| Genetic Risk Category, n (%) | Low PRS | 72741 (33.5) | 9848 (33.2) | 38095 (33.1) | 0.074 |
|  | Mid PRS | 72467 (33.4) | 9865 (33.3) | 38352 (33.3) |  |
|  | High PRS | 72048 (33.2) | 9946 (33.5) | 38688 (33.6) |  |
| FIB-4 Fibrosis Risk, n (%) | Low Risk | 124993 (57.5) | 14060 (47.4) | 64852 (56.3) | **<0.001** |
|  | Intermediate Risk | 88594 (40.8) | 14663 (49.4) | 47490 (41.2) |  |
|  | High Risk | 3669 (1.7) | 936 (3.2) | 2793 (2.4) |  |

Continuous variables are presented as means (standard deviations, SD), and categorical variables are presented as frequencies (percentages). Inter-group differences across the three Gaussian Mixture Model (GMM) clusters ("Healthy", " Fibro-Inflammatory", and " High-Risk Comorbid") were evaluated using Analysis of Variance (ANOVA) for continuous variables and Pearson's Chi-square test for categorical variables. **Abbreviations:** AF, atrial fibrillation; ALT, alanine aminotransferase; AST, aspartate aminotransferase; BMI, body mass index; FIB-4, Fibrosis-4 index; PRS, polygenic risk score.

#### Supplementary Table S9. Independent association between metabolic phenotypes and incident atrial fibrillation.

| **Model** | **Exposure** | **Type** | **Level** | **No.**  **(Total / Cases)** | **HR (95% CI)** | **P value** |
| --- | --- | --- | --- | --- | --- | --- |
| Crude (Unadjusted) | Metabolic Clusters | Categorical | Healthy (Ref) | 217256 / 11859 | 1.00 (Reference) | - |
|  |  |  | Fibro-Inflammatory | 29659 / 2110 | 1.34 (1.28-1.41) | **<0.001** |
|  |  |  | High-Risk Comorbid | 115135 / 10026 | 1.65 (1.61-1.70) | **<0.001** |
| Model 1 |  |  | Healthy (Ref) | 217256 / 11859 | 1.00 (Reference) | - |
|  |  |  | Fibro-Inflammatory | 29659 / 2110 | 1.24 (1.18-1.30) | **<0.001** |
|  |  |  | High-Risk Comorbid | 115135 / 10026 | 1.31 (1.27-1.34) | **<0.001** |
| Model 2 |  |  | Healthy (Ref) | 217256 / 11859 | 1.00 (Reference) | - |
|  |  |  | Fibro-Inflammatory | 29659 / 2110 | 1.25 (1.19-1.31) | **<0.001** |
|  |  |  | High-Risk Comorbid | 115135 / 10026 | 1.12 (1.08-1.15) | **<0.001** |
| Model 3 |  |  | Healthy (Ref) | 217256 / 11859 | 1.00 (Reference) | - |
|  |  |  | Fibro-Inflammatory | 29659 / 2110 | 1.19 (1.14-1.25) | **<0.001** |
|  |  |  | High-Risk Comorbid | 115135 / 10026 | 1.04 (1.01-1.07) | **0.013** |
| Model 4 |  |  | Healthy (Ref) | 217256 / 11859 | 1.00 (Reference) | - |
|  |  |  | Fibro-Inflammatory | 29659 / 2110 | 1.19 (1.14-1.25) | **<0.001** |
|  |  |  | High-Risk Comorbid | 115135 / 10026 | 1.04 (1.01-1.07) | **0.010** |

Hazard ratios (HRs) and 95% confidence intervals (CIs) were estimated using Cox proportional hazards regression models. For continuous analyses, liver fibrosis indices were winsorized at the 99th percentile and standardized; HRs reflect the risk per 1 standard deviation (SD) increase. Categorical groups were defined based on established clinical cutoffs, with the lowest risk group serving as the reference (Ref). **Crude:** Unadjusted model. **Model 1:** Adjusted for age, sex, and ethnicity. **Model 2:** Adjusted for Model 1 covariates plus body mass index, smoking status, alcohol intake frequency, and Townsend deprivation index. **Model 3:** Adjusted for Model 2 covariates plus hypertension, type 2 diabetes mellitus, lipid-lowering medication use, chronic kidney disease, and prevalent cardiovascular disease. **Model 4:** Adjusted for Model 3 covariates plus the standardized polygenic risk score (PRS) for atrial fibrillation and the first 10 genetic principal components. Boldface values indicate statistical significance (*P* < 0.05). **Abbreviations:** HR, Hazard ratio; CI, Confidence interval; Ref, Reference; AF, Atrial fibrillation.

#### Supplementary Table S10. Joint Effects of Genetic Predisposition and Metabolomic Risk on Incident Atrial Fibrillation.

| Cohort | Genetic Risk | Metabolic Risk | No. (Total/Cases) | HR (95% CI) | *P* Value |
| --- | --- | --- | --- | --- | --- |
| Full  (n = 362,050; 23,995 incident AF events) | Low PRS | Low MRS | 41689/1038 | 1.00 (Reference) | **-** |
|  |  | Mid MRS | 40147/1362 | 1.12 (1.04-1.22) | **0.005** |
|  |  | High MRS | 39150/2419 | 1.34 (1.24-1.44) | **<0.001** |
|  | Mid PRS | Low MRS | 39971/1536 | 1.56 (1.44-1.68) | **<0.001** |
|  |  | Mid MRS | 39901/2009 | 1.69 (1.57-1.82) | **<0.001** |
|  |  | High MRS | 40455/3767 | 2.06 (1.92-2.21) | **<0.001** |
|  | High PRS | Low MRS | 39027/2596 | 2.75 (2.56-2.96) | **<0.001** |
|  |  | Mid MRS | 40412/3512 | 3.02 (2.81-3.24) | **<0.001** |
|  |  | High MRS | 41298/5756 | 3.26 (3.04-3.49) | **<0.001** |
|  | Low PRS | Low MRS | 12587/295 | 1.00 (Reference) | **-** |
| Test  (n = 108,616; 7,031 incident AF events) |  | Mid MRS | 12151/425 | 1.22 (1.05-1.41) | **0.010** |
|  |  | High MRS | 11770/725 | 1.40 (1.22-1.61) | **<0.001** |
|  | Mid PRS | Low MRS | 11848/462 | 1.67 (1.44-1.93) | **<0.001** |
|  |  | Mid MRS | 11842/584 | 1.77 (1.54-2.04) | **<0.001** |
|  |  | High MRS | 12159/1091 | 2.13 (1.86-2.44) | **<0.001** |
|  | High PRS | Low MRS | 11774/742 | 2.74 (2.39-3.13) | **<0.001** |
|  |  | Mid MRS | 11989/1037 | 3.19 (2.80-3.64) | **<0.001** |
|  |  | High MRS | 12496/1670 | 3.28 (2.88-3.73) | **<0.001** |

Hazard ratios (HRs) and 95% confidence intervals (CIs) were estimated using Cox proportional hazards regression models pooled across 10 multiply imputed datasets via Rubin’s rules. Participants were stratified into nine joint categories based on tertiles of the polygenic risk score (PRS: low, mid, high) and the metabolomic risk score (MRS: low, mid, high). The stratum with the lowest genetic and metabolic risk (low PRS and low MRS) served as the universal reference category. The embedded visual forest plot represents the point estimates (diamonds) and 95% CIs (horizontal dashed lines) for the HRs across strata, with colors indicating the MRS risk levels. All models were comprehensively adjusted for baseline age, sex, ethnicity, body mass index, smoking status, alcohol intake frequency, Townsend deprivation index, hypertension, type 2 diabetes mellitus, lipid-lowering medication use, chronic kidney disease, prevalent cardiovascular disease, and the first 10 genetic principal components. **Abbreviations***:* CI, confidence interval; HR, hazard ratio; MRS, metabolomic risk score; PRS, polygenic risk score.

#### Supplementary Table S11. Additive interaction between the Metabolomic Risk Score (MRS) and Polygenic Risk Score (PRS) on the incidence of atrial fibrillation.

|  | Cohort | Metabolic Risk Level | Mid PRS | | High PRS | |
| --- | --- | --- | --- | --- | --- | --- |
|  |  |  | RERI (95% CI) | AP (95% CI) | RERI (95% CI) | AP (95% CI) |
|  | Full  (n = 362,050; 23,995 incident AF events) | Intermediate Risk | 0.01  (-0.13, 0.15) | 0.01 | 0.14  (-0.02, 0.31) | 0.05 |
|  |  | High Risk | 0.17  (0.04, 0.29) | 0.08 | 0.17  (0.01, 0.33) | 0.05 |
|  |  | *P* _multiplicative interaction_ < **0.001** | | | | |
| Test  (n = 108,616; 7,031 incident AF events) | | Intermediate Risk | -0.12  (-0.39, 0.15) | -0.07 | 0.24  (-0.08, 0.55) | 0.07 |
|  |  | High Risk | 0.06  (-0.19, 0.31) | 0.03 | 0.14  (-0.16, 0.44) | 0.04 |
|  |  | *P* _multiplicative interaction_ = **0.026** | | | | |

The table presents measures of additive interaction, including the Relative Excess Risk due to Interaction (RERI) and the Attributable Proportion (AP) with their 95% confidence intervals (CIs). The stratum with the lowest genetic and metabolic risk (Low PRS and Low MRS) served as the universal reference category. A RERI or AP significantly greater than 0 indicates a positive additive interaction (synergism), meaning that the combined effect of the genetic and metabolic risk factors is strictly larger than the sum of their individual effects.

#### Supplementary Table S12. Reclassification improvement of the Metabolomic Risk Score (MRS) for incident atrial fibrillation.

| **Comparison** | **Continuous NRI (95% CI)** | **NRI P-value** | **IDI (95% CI)** | **IDI P-value** |
| --- | --- | --- | --- | --- |
| 1. CHARGE-AF vs +MRS | 0.102 (0.050-0.149) | < 0.001 | 0.0014 (0.0010-0.0018) | < 0.001 |
| 2. CHARGE-AF+PRS vs +PRS+MRS | 0.093 (0.041-0.139) | < 0.001 | 0.0015 (0.0010-0.0020) | < 0.001 |
| 3. ARIC vs +MRS | 0.117 (0.066-0.168) | < 0.001 | 0.0012 (0.0008-0.0016) | < 0.001 |
| 4. ARIC+PRS vs +PRS+MRS | 0.110 (0.058-0.160) | < 0.001 | 0.0013 (0.0008-0.0018) | < 0.001 |
| 5. C2HEST vs +MRS | 0.322 (0.272-0.367) | < 0.001 | 0.0031 (0.0026-0.0035) | < 0.001 |
| 6. C2HEST+PRS vs +PRS+MRS | 0.314 (0.268-0.362) | < 0.001 | 0.0040 (0.0034-0.0046) | < 0.001 |

Continuous Net Reclassification Improvement (cNRI) and Integrated Discrimination Improvement (IDI) evaluate the incremental predictive value of adding the MRS to established clinical models (CHARGE-AF, ARIC, and C2HEST), with or without the Polygenic Risk Score (PRS). Reclassification metrics were assessed at a [5-year] follow-up horizon in the independent test cohort. The 95% confidence intervals (CIs) for cNRI were derived from 500 bootstrap iterations. **Abbreviations:** AF, atrial fibrillation; CI, confidence interval; cNRI, continuous net reclassification improvement; IDI, integrated discrimination improvement; MRS, Metabolomic Risk Score; PRS, polygenic risk score.

#### Supplementary Table S13. Missing data rates across the four study cohorts.

| **Variable** | **Raw Population (Before Exclusion)** | | | | **Analytical Cohort (After Exclusion)** | | | |
| --- | --- | --- | --- | --- | --- | --- | --- | --- |
|  | **Total N** | **Valid N** | **Missing N** | **Missing**  **(%)** | **Total N** | **Valid N** | **Missing N** | **Missing**  **(%)** |
| **Cohort: Main Cohort (Baseline)** | | | | | | | | |
| Systolic blood pressure (reading 2) | 501,911 | 460,754 | 41,157 | 8.20 | 403,974 | 372,396 | 31,578 | 7.82 |
| Diastolic blood pressure (reading 2) | 501,911 | 460,758 | 41,153 | 8.20 | 403,974 | 372,398 | 31,576 | 7.82 |
| Systolic blood pressure (reading 1) | 501,911 | 467,524 | 34,387 | 6.85 | 403,974 | 377,956 | 26,018 | 6.44 |
| Diastolic blood pressure (reading 1) | 501,911 | 467,537 | 34,374 | 6.85 | 403,974 | 377,967 | 26,007 | 6.44 |
| Glycated haemoglobin (HbA1c) | 501,911 | 465,985 | 35,926 | 7.16 | 403,974 | 384,709 | 19,265 | 4.77 |
| Glucose | 501,911 | 429,090 | 72,821 | 14.51 | 403,974 | 403,366 | 608 | 0.15 |
| Townsend deprivation index | 501,911 | 501,290 | 621 | 0.12 | 403,974 | 403,473 | 501 | 0.12 |
| Ethnic background | 501,911 | 501,022 | 889 | 0.18 | 403,974 | 403,584 | 390 | 0.10 |
| Smoking status | 501,911 | 501,029 | 882 | 0.18 | 403,974 | 403,586 | 388 | 0.10 |
| HDL cholesterol | 501,911 | 429,393 | 72,518 | 14.45 | 403,974 | 403,669 | 305 | 0.08 |
| Triglycerides | 501,911 | 468,688 | 33,223 | 6.62 | 403,974 | 403,673 | 301 | 0.07 |
| Creatinine | 501,911 | 468,831 | 33,080 | 6.59 | 403,974 | 403,742 | 232 | 0.06 |
| Gamma glutamyltransferase | 501,911 | 468,815 | 33,096 | 6.59 | 403,974 | 403,764 | 210 | 0.05 |
| Waist circumference | 501,911 | 499,775 | 2,136 | 0.43 | 403,974 | 403,899 | 75 | 0.02 |
| Age | 501,911 | 501,911 | 0 | 0.00 | 403,974 | 403,974 | 0 | 0.00 |
| Sex | 501,911 | 501,911 | 0 | 0.00 | 403,974 | 403,974 | 0 | 0.00 |
| Body mass index (BMI) | 501,911 | 498,831 | 3,080 | 0.61 | 403,974 | 403,974 | 0 | 0.00 |
| Alanine aminotransferase | 501,911 | 468,874 | 33,037 | 6.58 | 403,974 | 403,974 | 0 | 0.00 |
| Aspartate aminotransferase | 501,911 | 467,271 | 34,640 | 6.90 | 403,974 | 403,974 | 0 | 0.00 |
| Platelet count | 501,911 | 477,635 | 24,276 | 4.84 | 403,974 | 403,974 | 0 | 0.00 |
| Albumin | 501,911 | 429,597 | 72,314 | 14.41 | 403,974 | 403,974 | 0 | 0.00 |
| Standard PRS for atrial fibrillation (AF) | 501,911 | 485,717 | 16,194 | 3.23 | 403,974 | 403,974 | 0 | 0.00 |
| **NMR Multiomics Cohort (Baseline)** | | | | | | | | |
| Complete NMR Metabolomics Profile | 501,911 | 441,492 | 60,419 | 12.04 | 362,050 | 362,050 | 0 | 0.00 |
| **MRI Subcohort (Instance 2)** | | | | | | | | |
| Systolic blood pressure (reading 2) | 27,637 | 20,690 | 6,947 | 25.14 | 27,032 | 20,220 | 6,812 | 25.20 |
| Diastolic blood pressure (reading 2) | 27,637 | 20,693 | 6,944 | 25.13 | 27,032 | 20,223 | 6,809 | 25.19 |
| Systolic blood pressure (reading 1) | 27,637 | 21,149 | 6,488 | 23.48 | 27,032 | 20,660 | 6,372 | 23.57 |
| Diastolic blood pressure (reading 1) | 27,637 | 21,149 | 6,488 | 23.48 | 27,032 | 20,660 | 6,372 | 23.57 |
| Waist circumference | 27,637 | 26,610 | 1,027 | 3.72 | 27,032 | 26,023 | 1,009 | 3.73 |
| Proton density fat fraction (PDFF) | 27,637 | 27,312 | 325 | 1.18 | 27,032 | 26,717 | 315 | 1.17 |
| Smoking status | 27,637 | 27,477 | 160 | 0.58 | 27,032 | 26,880 | 152 | 0.56 |
| Liver iron corrected T1 (ct1) | 27,637 | 27,637 | 0 | 0.00 | 27,032 | 27,032 | 0 | 0.00 |
| Alcohol drinker status | 27,637 | 27,637 | 0 | 0.00 | 27,032 | 27,032 | 0 | 0.00 |
| Alcohol intake frequency. | 27,637 | 27,637 | 0 | 0.00 | 27,032 | 27,032 | 0 | 0.00 |
| **Cardiac Phenotype Subcohorts (Instance 2)** | | | | | | | | |
| Left atrial volume index (LAVI) | 501,911 | 38,011 | 463,900 | 92.43 | 26,426 | 26,426 | 0 | 0 |
| LV global longitudinal strain (GLS) | 501,911 | 38,091 | 463,820 | 92.41 | 26,532 | 26,532 | 0 | 0 |
| Left ventricular mass index (LVMI) | 501,911 | 38,542 | 463,369 | 92.32 | 26,779 | 26,779 | 0 | 0 |
| LV end-diastolic volume index (LVEDVI) | 501,911 | 38,542 | 463,369 | 92.32 | 26,779 | 26,779 | 0 | 0 |
| Left atrial active emptying fraction (LAAEF) | 501,911 | 38,814 | 463,097 | 92.27 | 26,995 | 26,995 | 0 | 0 |
| Left ventricular ejection fraction (LVEF) | 501,911 | 39,565 | 462,346 | 92.12 | 27,480 | 27,480 | 0 | 0 |
| PQ interval | 501,911 | 43,895 | 458,016 | 91.25 | 30,529 | 30,529 | 0 | 0 |
| QTC interval | 501,911 | 46,392 | 455,519 | 90.76 | 31,990 | 31,990 | 0 | 0 |
| P duration | 501,911 | 76,847 | 425,064 | 84.69 | 53,837 | 53,837 | 0 | 0 |
| QRS duration | 501,911 | 81,369 | 420,542 | 83.79 | 56,348 | 56,348 | 0 | 0 |
| Ventricular rate | 501,911 | 81,379 | 420,532 | 83.79 | 56,353 | 56,353 | 0 | 0 |

The 'Raw Population' represents all available UK Biobank participants at the respective baseline or imaging visits prior to exclusions. The 'Analytical Cohort' denotes the final participant subsets retained for statistical modeling after applying strict exclusion criteria (e.g., prevalent atrial fibrillation and missing core variates). Notably, the 'Complete NMR Metabolomics Profile' treats the concurrent availability of all 249 quantitative metabolic features as a single composite analytical unit. **Abbreviations:** AF, atrial fibrillation; BMI, body mass index; cT1, iron-corrected T1; GLS, global longitudinal strain; HbA1c, glycated haemoglobin; LAAEF, left atrial active emptying fraction; LAVI, left atrial volume index; LVEDVI, left ventricular end-diastolic volume index; LVEF, left ventricular ejection fraction; LVMI, left ventricular mass index; MRI, magnetic resonance imaging; NMR, nuclear magnetic resonance; PDFF, proton density fat fraction; PRS, polygenic risk score; RAVI, right atrial volume index.

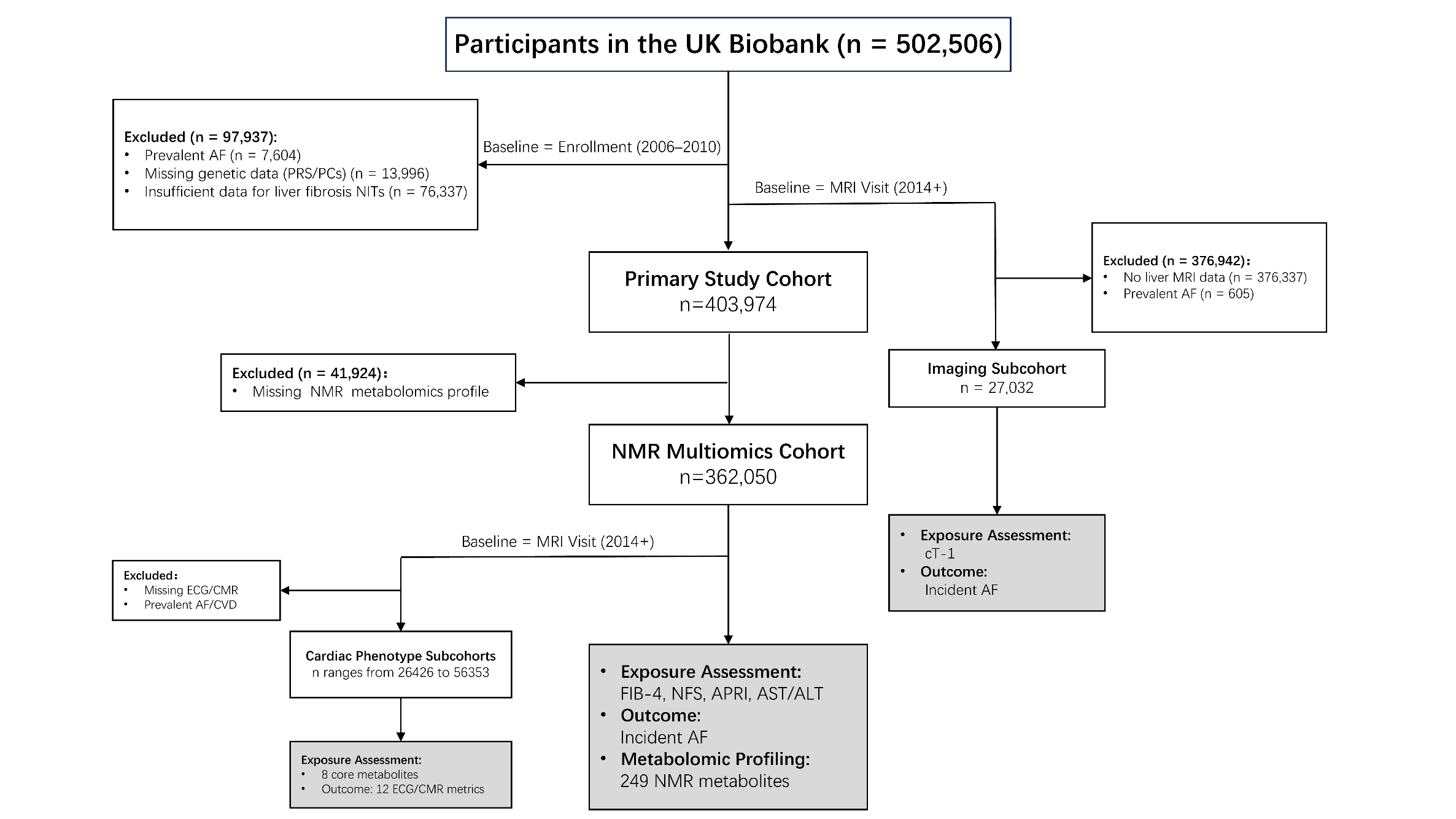

#### Supplementary Figure S1. Flowchart of study participant selection and subcohort definitions.

#### The flowchart details the stepwise inclusion and exclusion criteria for the primary analytical cohort (n = 403,974) and three deeply phenotyped subcohorts derived from the UK Biobank. The Imaging Subcohort (n = 27,032), NMR Multi-omics Cohort (n = 362,050), and Cardiac Phenotype Subcohorts (n = 26426 to 56353) were established by applying strict data availability thresholds and disease-free criteria to rigorously evaluate subclinical remodeling and metabolomic profiles. Abbreviations: AF, atrial fibrillation; PRS, polygenic risk score; PCs, principal components; NITs, noninvasive tests; cT1, corrected T1; NMR, nuclear magnetic resonance; LASSO, least absolute shrinkage and selection operator; ECG, electrocardiogram; CMR, cardiac magnetic resonance; CVD, cardiovascular disease.

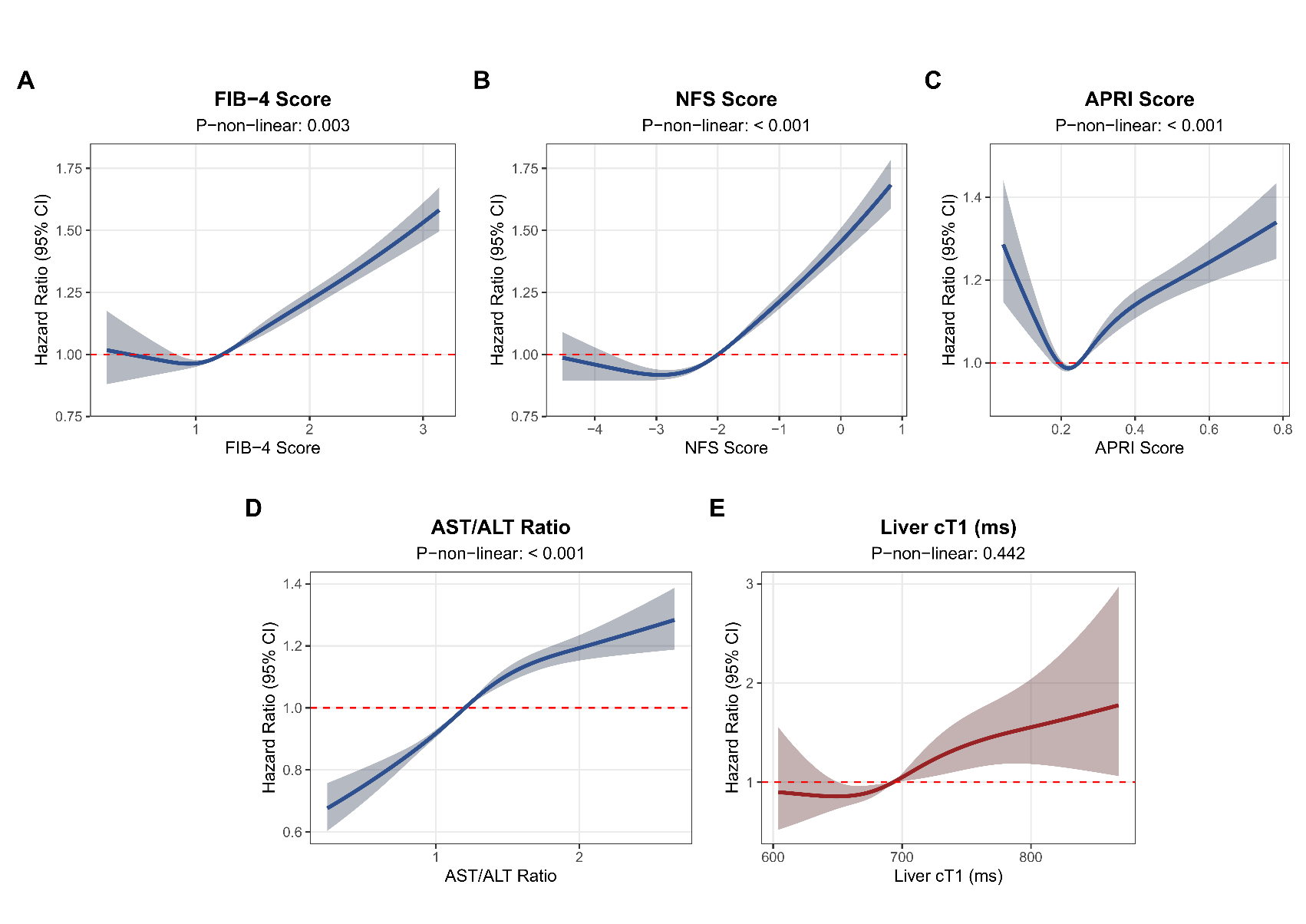

#### Supplementary Figure S2. Dose-response associations of liver fibrosis indices and liver cT1 with incident atrial fibrillation.

Restricted cubic splines (RCS, 4 knots) display the multivariable-adjusted hazard ratios (solid lines) and 95% confidence intervals (shaded areas) for **(A)** FIB-4, **(B)** NFS, **(C)** APRI, **(D)** AST/ALT ratio, and **(E)** Liver cT1. The reference point (HR = 1.0, red dashed line) was set at the median value of each index after winsorization (1st–99th percentiles for NFS and cT1; up to 99th percentile for others). Analyses for panels A–D were based on the fully adjusted Model 4 in the main cohort, while panel E was based on the fully adjusted Model 4 in the MRI subcohort. Nonlinearity was evaluated using the likelihood ratio test ($P_{\text{non-linear}}$). **Abbreviations:** ALT, alanine aminotransferase; APRI, AST to platelet ratio index; AST, aspartate aminotransferase; CI, confidence interval; cT1, corrected T1; FIB-4, Fibrosis-4 index; HR, hazard ratio; NFS, nonalcoholic fatty liver disease (NAFLD) fibrosis score; RCS, restricted cubic splines.

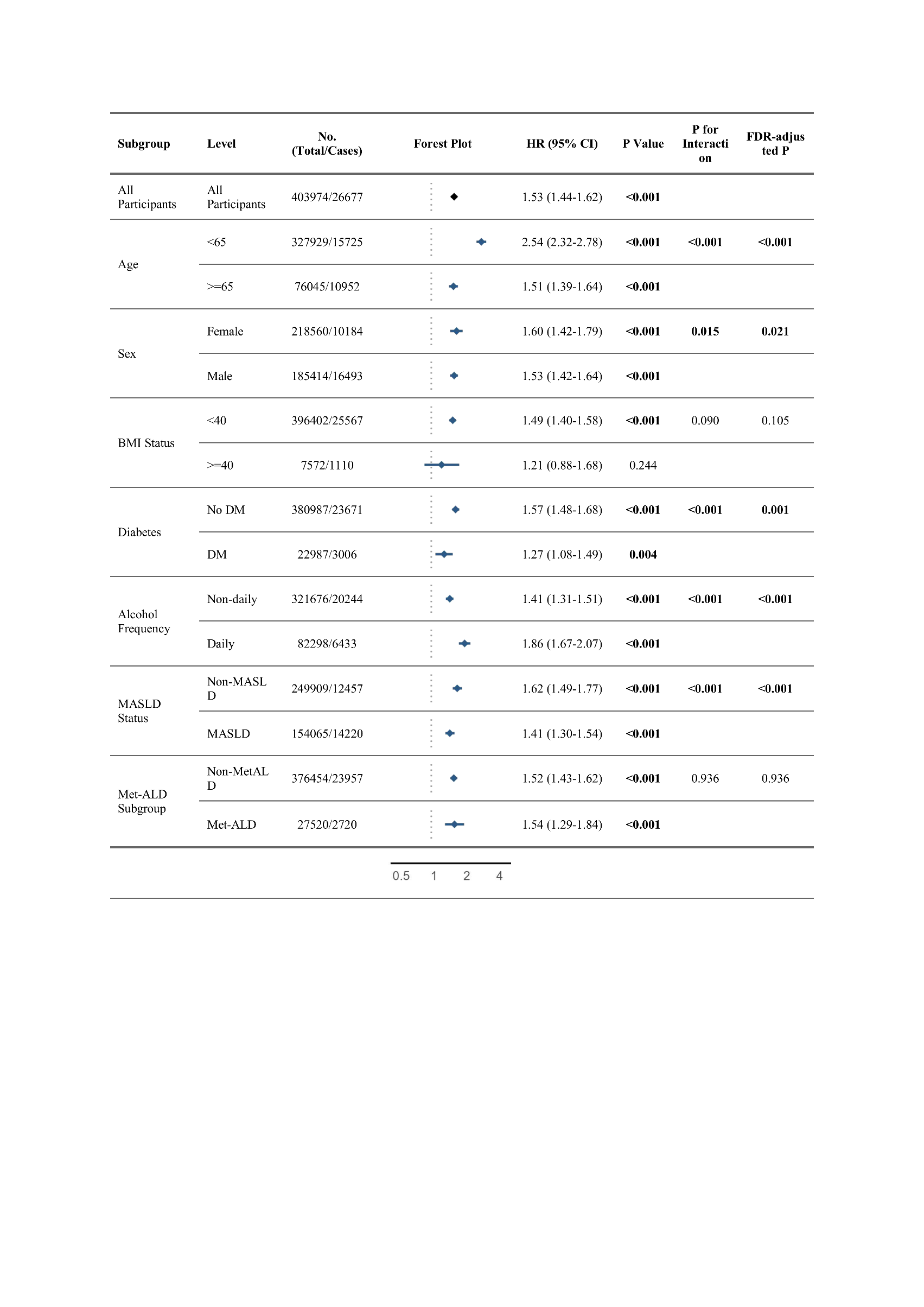

#### Supplementary Figure S3. Subgroup analyses for the association between high-risk FIB-4 index and incident atrial fibrillation.

The integrated forest plot and table illustrate the hazard ratios (HRs, represented by diamonds) and 95% confidence intervals (CIs, represented by horizontal solid lines) for incident atrial fibrillation (AF) associated with a high-risk FIB-4 index across pre-specified clinical subgroups. The vertical dotted line indicates an HR of 1.0 (no effect). Hazard ratios and interaction *P*-values were estimated using multivariable Cox proportional hazards models pooled across 10 multiply imputed datasets via Rubin's rules. Hazard ratios were derived from the fully adjusted Model 4 (covariates are detailed in the main Methods section). To account for multiple testing across subgroups, P-values for interaction ($P_{\text{interaction}}$) were adjusted using the False Discovery Rate (FDR) method via the Benjamini-Hochberg procedure. Within-subgroup hazard ratios and their associated P-values are presented without multiple-testing correction to reflect primary stratum-specific estimates. **Note:** To strictly prevent multicollinearity, the specific stratifying variable was systematically omitted from the adjustment covariates in its respective subgroup model. **Abbreviations:** AF, atrial fibrillation; CI, confidence interval; HR, hazard ratio; BMI, body mass index; DM, diabetes mellitus; MASLD, metabolic dysfunction-associated steatotic liver disease; MetALD, metabolic dysfunction and alcohol-associated steatotic liver disease; FIB-4, Fibrosis-4 index; FDR, false discovery rate.

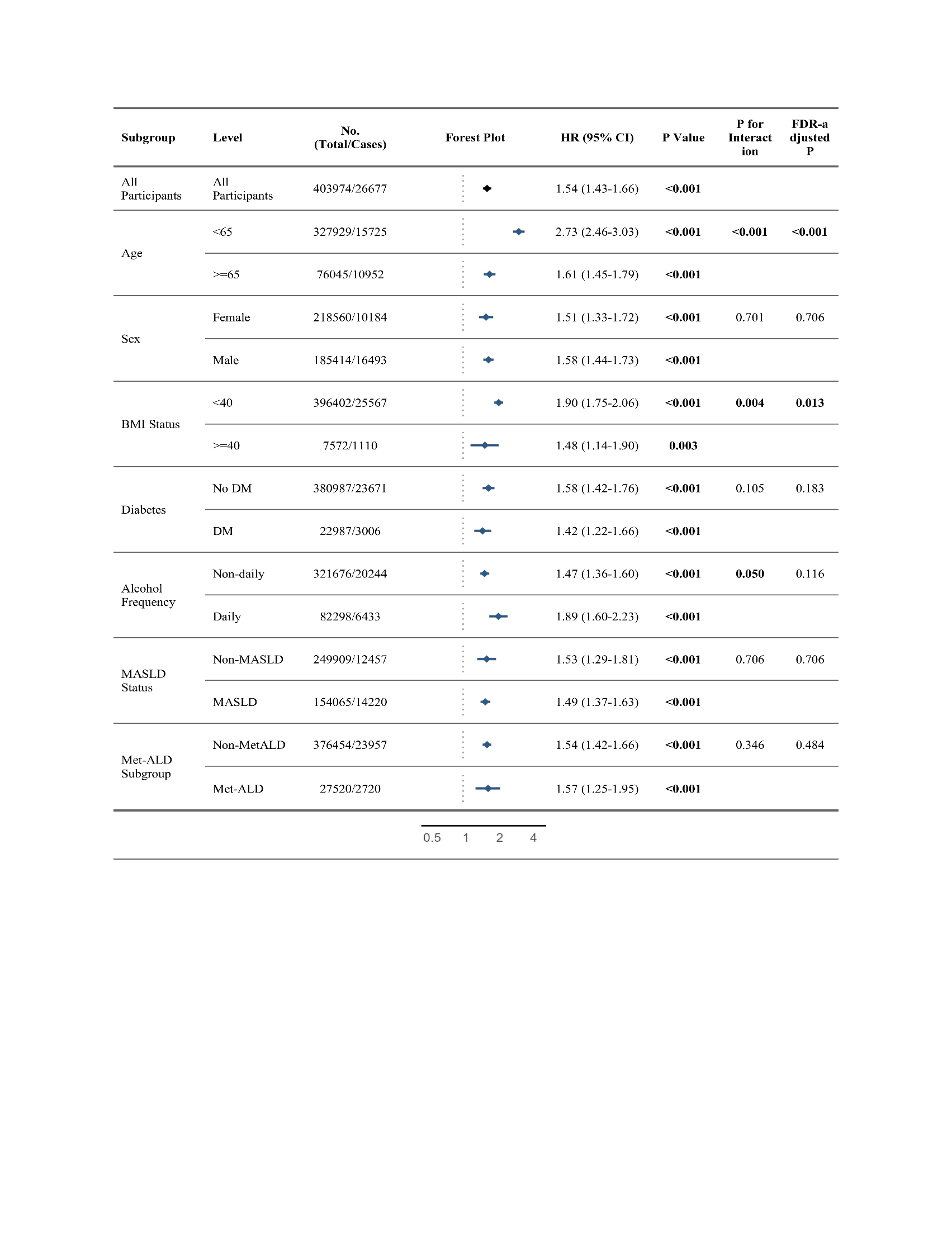

#### Supplementary Figure S4. Subgroup analyses for the association between high-risk NFS and incident atrial fibrillation.

The integrated forest plot and table illustrate the hazard ratios (HRs, represented by diamonds) and 95% confidence intervals (CIs, represented by horizontal solid lines) for incident atrial fibrillation (AF) associated with a high-risk NFS across pre-specified clinical subgroups. The vertical dotted line indicates an HR of 1.0 (no effect). Hazard ratios and interaction *P*-values were estimated using multivariable Cox proportional hazards models pooled across 10 multiply imputed datasets via Rubin's rules. Hazard ratios were derived from the fully adjusted Model 4 (covariates are detailed in the main Methods section). To account for multiple testing across subgroups, P-values for interaction ($P_{\text{interaction}}$) were adjusted using the False Discovery Rate (FDR) method via the Benjamini-Hochberg procedure. Within-subgroup hazard ratios and their associated P-values are presented without multiple-testing correction to reflect primary stratum-specific estimates. **Note:** To strictly prevent multicollinearity, the specific stratifying variable was systematically omitted from the adjustment covariates in its respective subgroup model. **Abbreviations:** AF, atrial fibrillation; CI, confidence interval; HR, hazard ratio; BMI, body mass index; DM, diabetes mellitus; MASLD, metabolic dysfunction-associated steatotic liver disease; MetALD, metabolic dysfunction and alcohol-associated steatotic liver disease; NFS, NAFLD fibrosis score; FDR, false discovery rate.

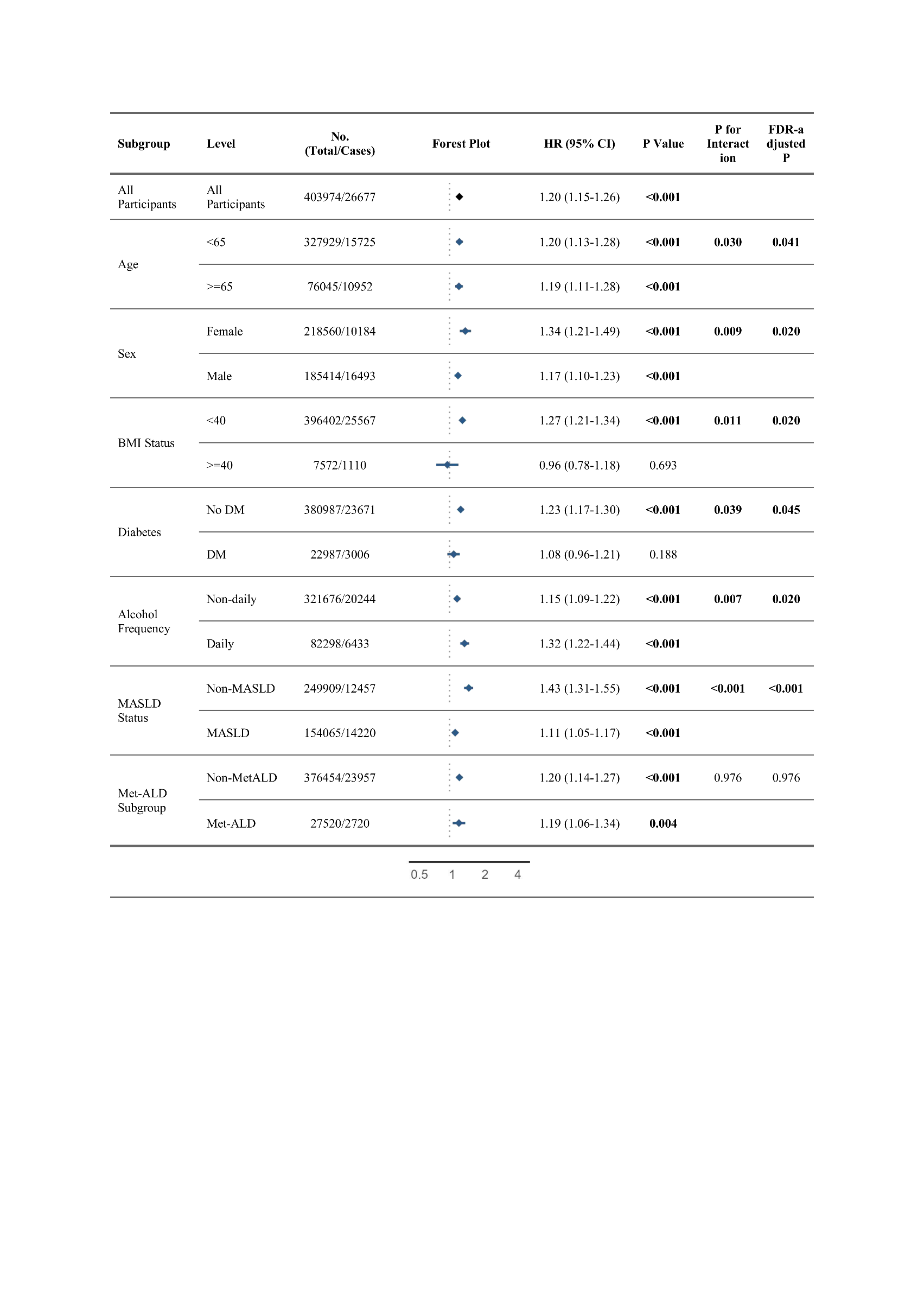

#### Supplementary Figure S5. Subgroup analyses for the association between high-risk APRI and incident atrial fibrillation.

The integrated forest plot and table illustrate the hazard ratios (HRs, represented by diamonds) and 95% confidence intervals (CIs, represented by horizontal solid lines) for incident atrial fibrillation (AF) associated with a high-risk APRI across pre-specified clinical subgroups. The vertical dotted line indicates an HR of 1.0 (no effect). Hazard ratios and interaction *P*-values were estimated using multivariable Cox proportional hazards models pooled across 10 multiply imputed datasets via Rubin's rules. Hazard ratios were derived from the fully adjusted Model 4 (covariates are detailed in the main Methods section). To account for multiple testing across subgroups, P-values for interaction ($P_{\text{interaction}}$) were adjusted using the False Discovery Rate (FDR) method via the Benjamini-Hochberg procedure. Within-subgroup hazard ratios and their associated P-values are presented without multiple-testing correction to reflect primary stratum-specific estimates. **Note:** To strictly prevent multicollinearity, the specific stratifying variable was systematically omitted from the adjustment covariates in its respective subgroup model. **Abbreviations:** AF, atrial fibrillation; CI, confidence interval; HR, hazard ratio; BMI, body mass index; DM, diabetes mellitus; MASLD, metabolic dysfunction-associated steatotic liver disease; MetALD, metabolic dysfunction and alcohol-associated steatotic liver disease; APRI, AST to Platelet Ratio Index; FDR, false discovery rate.

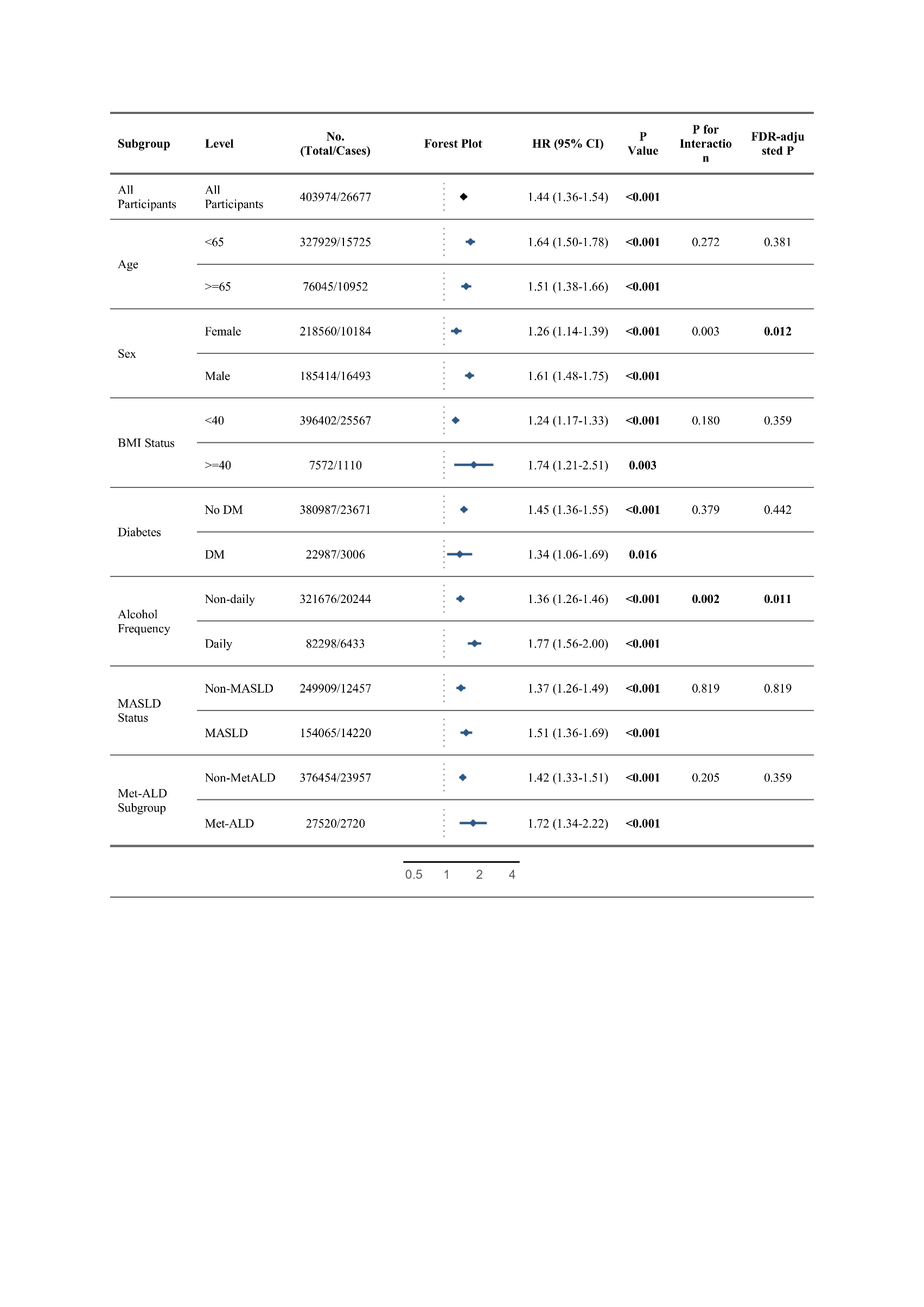

#### Supplementary Figure S6. Subgroup analyses for the association between high-risk AST/ALT ratio and incident atrial fibrillation.

The integrated forest plot and table illustrate the hazard ratios (HRs, represented by diamonds) and 95% confidence intervals (CIs, represented by horizontal solid lines) for incident atrial fibrillation (AF) associated with a high-risk AST/ALT ratio across pre-specified clinical subgroups. The vertical dotted line indicates an HR of 1.0 (no effect). Hazard ratios and interaction *P*-values were estimated using multivariable Cox proportional hazards models pooled across 10 multiply imputed datasets via Rubin's rules. Hazard ratios were derived from the fully adjusted Model 4 (covariates are detailed in the main Methods section). To account for multiple testing across subgroups, P-values for interaction ($P_{\text{interaction}}$) were adjusted using the False Discovery Rate (FDR) method via the Benjamini-Hochberg procedure. Within-subgroup hazard ratios and their associated P-values are presented without multiple-testing correction to reflect primary stratum-specific estimates. **Note:** To strictly prevent multicollinearity, the specific stratifying variable was systematically omitted from the adjustment covariates in its respective subgroup model. **Abbreviations:** AF, atrial fibrillation; CI, confidence interval; HR, hazard ratio; BMI, body mass index; DM, diabetes mellitus; MASLD, metabolic dysfunction-associated steatotic liver disease; MetALD, metabolic dysfunction and alcohol-associated steatotic liver disease; AST, aspartate aminotransferase; ALT, alanine transaminase; FDR, false discovery rate.

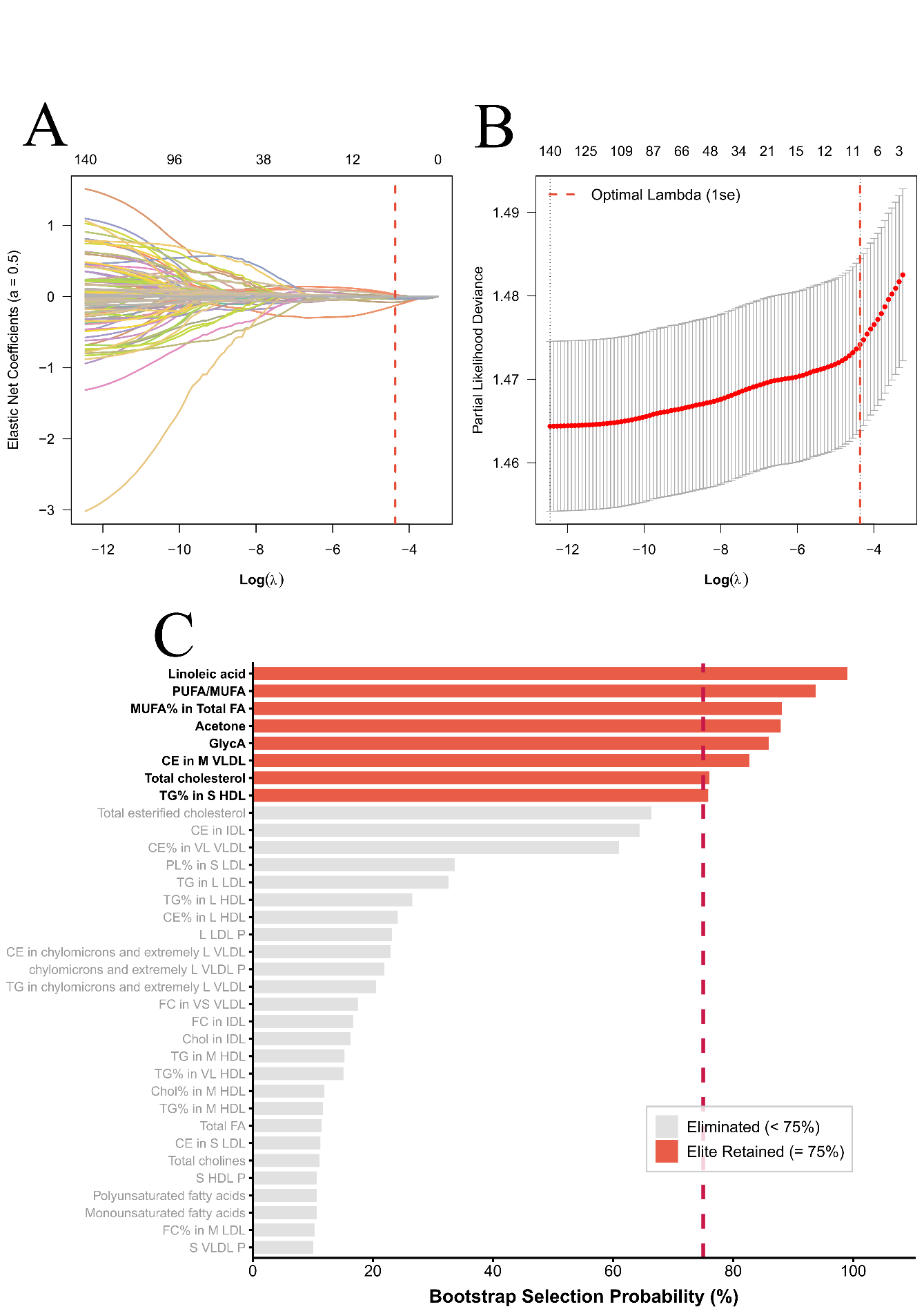

#### Supplementary Figure S7. Identification of ultra-stable mediators via Elastic Net penalized regression and bootstrap stability selection.

**(A)** Elastic Net coefficient paths. The coefficient paths of the 144 core metabolites are plotted against the $\log\left( \lambda\right)$ sequence ($\alpha= 0.5$). The top axis indicates the number of non-zero coefficients. The vertical red dashed line indicates the optimal $\lambda$ selected by the 1-standard error (1-se) rule. **(B)** Cross-validation curve. Ten-fold cross-validation error (partial likelihood deviance) across the $\log\left( \lambda\right)$ sequence. Error bars represent standard deviations. The optimal 1-se $\lambda$ is marked by the dashed red line. **(C)** Bootstrap stability selection. Bar chart displaying the selection probability of metabolites across 500 bootstrap iterations. Only metabolites with a selection frequency > 10% are shown. The vertical dashed line demarcates the 75% threshold used to define the final "ultra-stable" elite mediators (highlighted in red). **Abbreviations:** PUFA, Polyunsaturated fatty acids; MUFA, Monounsaturated fatty acids; FA, Fatty acids; CE, Cholesteryl esters; FC, Free cholesterol; Chol, Cholesterol; PL, Phospholipids; TG, Triglycerides; GlycA, Glycoprotein acetyls; VLDL, Very low-density lipoprotein; LDL, Low-density lipoprotein; HDL, High-density lipoprotein; IDL, Intermediate-density lipoprotein; P, Particles; VL/L/M/S/VS, Very large/Large/Medium/Small/Very small particle size prefixes.

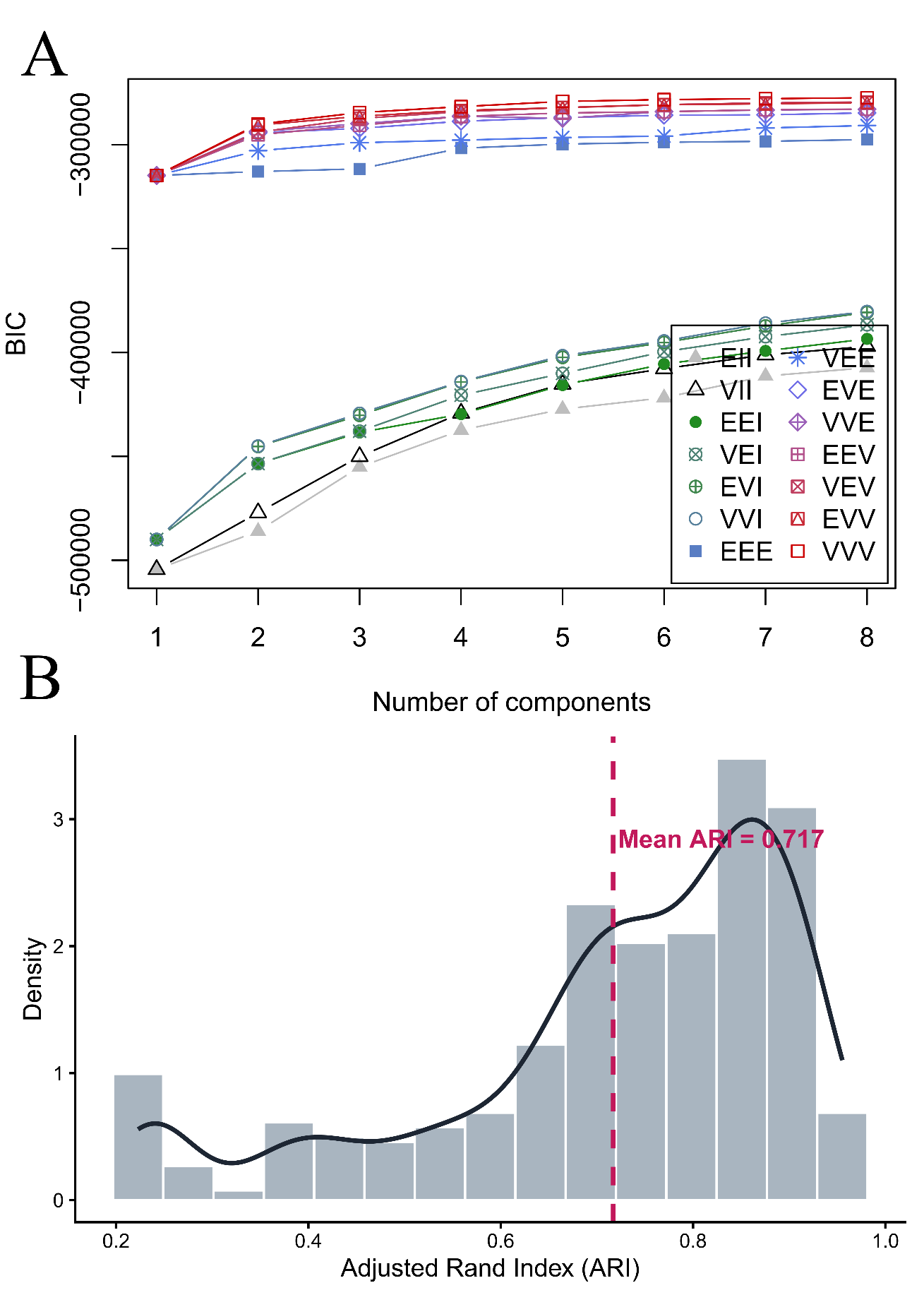

#### Supplementary Figure S8. Model selection and internal validation of Gaussian Mixture Model (GMM) clustering.

**(A)** Bayesian Information Criterion (BIC) curves. BIC values were evaluated for 1 to 8 mixture components across 14 parameterized covariance structures (indicated by different colors and symbols) using the ultra-stable core metabolites in the discovery cohort (70% split). **(B)** Bootstrap clustering stability. Histogram and density plot showing the distribution of the Adjusted Rand Index (ARI) calculated across 500 bootstrap iterations.
